# Pathogenesis of Multiple Sclerosis: Genetic, Environmental, and Random Mechanisms

**DOI:** 10.1101/2023.12.25.23300524

**Authors:** Douglas S. Goodin

**Affiliations:** Department of Neurology, University of California, San Francisco & the San Francisco VA Medical Center, San Francisco, CA, USA

**Keywords:** Multiple Sclerosis, pathogenesis, environmental factors, genetic factors, randomness, determinism

## Abstract

**BACKGROUND:** MS-pathogenesis requires both genetic factors and environmental events. The question remains, however, whether these factors and events *completely* describe the MS disease-process. This question was addressed using the Canadian MS-data, which includes 29,478 individuals, representing 65-83% of all Canadian MS-patients.

**METHODS:** The “*genetically-susceptible*” subset of the population, (*G*), includes *everyone* who has *any non-zero* life-time chance of developing MS, under *some* environmental-conditions. A “*sufficient*” environmental-exposure, for *any* “*genetically-susceptible*” individual, includes *every* set of environmental conditions, each of which is *sufficient*, by itself, to *cause* MS in that person. This analysis incorporates several different *epidemiologic-parameters*, involved in MS-pathogenesis, only some of which are directly-observable, and establishes “*plausible-value-ranges”* for each parameter. Those *parameter-value* combinations (solutions) that fall within these *plausible-ranges* are then determined.

**RESULTS:** Only a fraction of the population can possibly be “*genetically-susceptible*”. Thus, many individuals have *no possibility* of developing MS under *any* environmental conditions. Moreover, *some* “*genetically-susceptible*” individuals, despite their experiencing a “*sufficient*” environmental-exposure, *will never* develop disease.

**CONCLUSIONS:** This analysis *explicitly includes* all of those genetic factors and environmental events (including interactions), which are necessary for MS-pathogenesis, regardless of whether these are known, suspected, or as yet unrecognized. Nevertheless, in addition, “*true*” randomness seems to play a critical role in disease-pathogenesis. This observation provides empirical evidence that undermines the widely-held deterministic view of nature. Moreover, both sexes seem to have a similar genetic and environmental disease-basis. If so, this indicates that this random element is primarily responsible for the currently-observed differences in disease-expression between *susceptible-women* and *susceptible-men*.

## Introduction

Multiple sclerosis (MS)-pathogenesis requires both environmental-events and genetic-factors [1-4]. Considering genetics, familial-aggregation of MS-cases is well-established. MS-risk is increased ∼30-fold in non-twin siblings and ∼250-fold in monozygotic (*MZ*)-twins of an *MS-proband* [1,2,5]. Moreover, 233 MS-associated genetic-traits are now identified [6]. Nevertheless, MS-genetics is complex. The strongest MS-association is with the *Class-II* haplotype, *HLA-DRB1*15:01∼DQB1*06:02*, located at (6p21), having an odds-ratio (*OR*) of (∼3) in heterozygotes and of (∼6) in homozygotes [1,2,5,6]. Other MS-associations are quite weak [6] – (median − OR = 1.158; interquartile − range = 1.080 − 1.414). Also, *DRB1*15:01∼DQB1*06:02* is highly “selected”, accounting for (∼13%) of *DRB1∼DQB1-* haplotypes – the most frequent such haplotype – among European-decedents [1-8]. Moreover, *everyone* (except *MZ-*twins) possesses a unique combination of the 233 MS-associated genetic-traits [3]. Finally, the observed *probability-range* for *MZ-*twin-concordance is: (0.11 – 0.46) and, consequently, genetics plays only a minor role in determining disease-expression (*Table-4; Reference:[3]*).

MS is also linked to environmental-events. First, a well-documented *month-of-birth* effect, linking MS-risk to the solar cycle, likely implicates intrauterine/perinatal environmental-events in MS-pathogenesis [2,9-11]. Second, given an *MS-proband*, the MS-recurrence-rate for dizygotic (*DZ*)-twins exceeds that for non-twin siblings [2,3,5] – also implicating intrauterine/perinatal environmental-events [2,3]. Third, MS becomes increasingly prevalent farther north or south from equatorial-regions [2,12]. Because this gradient is also evident for *MZ-*twin-recurrence-rates (*Table-4; Reference:[3]*), environmental-factors are likely responsible. Fourth, a prior Epstein-Barr viral (*EBV*) infection is found in almost all (> 99%) *current* MS patients [2,13,14]. If these rare *EBV-negative* patients represent *false-negative-tests* – either from inherent-errors when using *any* fixed antibody-titer “*cut-off*” to determine *EBV-positivity*, or from *only* determining antibody-responses to *some EBV-antigens* [2] – then one can conclude that *EBV-*infection is a necessary-factor in *every* causal-pathway, which led to MS in these individuals [2]. Regardless, an *EBV*-infection *must* somehow be involved in MS-pathogenesis [2,13,14]. Lastly, smoking and vitamin-D deficiency are implicated in MS-pathogenesis [2,15,16].

This manuscript presents an analysis regarding genetic and environmental susceptibility to MS [4] in a relatively non-mathematical format to make its conclusions accessible. The terms and definitions used for this analysis are presented in *Table-1*. For interested readers, the mathematical-development is presented in the *Supplemental-Material*. This analysis is based on the *Canadian-Collaborative-Project-on-Genetic-Susceptibility-to-Multiple-Sclerosis* (*CCPGSMS*)-dataset [5,8,9,17-23] – a summary of which is provided in the *Supplemental-Material-Sections:10a-b*. The *CCPGSMS*-dataset includes 29,478 MS-patients (born: 1891 - 1993) – estimated to represent (65 - 83%) of Canadian MS-patients [5,23,24]. This cohort is assumed to represent a large random sample of the Canadian MS-population. Also, this single population provides point-estimates and confidence-intervals for the MS-recurrence-rates in *MZ*-twins, *DZ*-twins, and non-twin siblings (*S*), and for the time-dependent changes in the *female-to-male* (*F:M*)-*sex-ratio*.

**Table 1.** Definition of Terms.

| Terms | Definitions |
| --- | --- |
| $(Z)$ | The population – a set consisting of $(N)$ individuals |
| $(F), (M)$ | Subsets of *women* $(F)$ and *men* $(M)$ within $(Z)$ |
| $(MS)$ | Subset of *all* individuals within $(Z)$ who either have, or will subsequently develop, MS |
| $(G)$ | Subset of *all* individuals within $(Z)$ who have *any* non-zero chance of developing MS under *some* environmental conditions |
| $(MZ), (DZ), (S)$ | Subsets of monozygotic-twins $(MZ)$, dizygotic-twins $(DZ)$, and non-twin siblings $(S)$ within $(Z)$ |
| *Proband* | An individual, randomly-selected either from $(Z)$ or from one of its subsets |
| *Co-twin, Co-sibling* | Either a twin – $(MZ)$ or $(DZ)$ – or a non-twin sibling $(S)$ of the *proband* |
| *Recurrence Rate* | Probability that the *proband* is a member of the $(MS)$-subset, given that their *co-twin* or non-twin *co-sibling* is a member of the $(MS)$-subset |
| *Penetrance* | Probability that a *proband* will develop MS over the course of their *life-time* |
| $(MZ, MS)$ | Subset of *MZ-twin probands* within the $(MS)$ subset |
| $(MZ_{MS})$ | Subset of *MZ co-twins* within the $(MS)$ subset |
| $(DZ_{MS}), (S_{MS})$ | Subsets of *DZ co-twins* $(DZ_{MS})$ and non-twin *co-siblings* $(S_{MS})$ within the $(MS)$ subset |
| $P(MS \vert IG_{MS})$ | Recurrence-rate for *MZ-twins*, adjusted because *MZ-twins*, who share identical genotypes $(IG)$, also share their intrauterine and *some* of their childhood environments – this adjustment isolates the genetic contribution to *MZ-twin* recurrence-rates – *see Text* |
| $(E_T)$ | Some specific *Time-Period* – *see legend; Table 2* |
| $P(MS \vert E_T)$ | *Penetrance* of MS for the population $(Z)$ during $(E_T)$ – *see Text* |
| $C$ | Ratio of *MS-penetrance* during *Time-Period #1*, $P(MS)_1$, to that during *Time-Period #2*, $P(MS)_2$ |
| $P(MS \vert G, E_T)$ | *Penetrance* of MS for the $(G)$ subset of the population $(Z)$ during $(E_T)$ – *see Text* |
| $Zw$ | *Penetrance* of MS for the subset of *susceptible-women* $(F, G)$ within $(Z)$ during $(E_T)$<br>– Also called the *failure probability* for *susceptible-women* during $(E_T)$ |
| $Zm$ | *Penetrance* of MS for the subset of *susceptible-men* $(M, G)$ within $(Z)$ during $(E_T)$<br>– Also called the *failure probability* for *susceptible-men* during $(E_T)$ |
| $c, d$ | Limiting values (constants) for the *failure probability* in *susceptible-men* ($c$); and *susceptible-women* ($d$) – i.e., $(Zm \leq c \leq 1)$ and $(Zw \leq d \leq 1)$ |
| $p$ | Proportion of women in the $(G)$ subset – i.e., $p = P(F \vert G)$ |
| $(E)$ | Event that a randomly-selected member of $(G)$ – the *proband* – experiences an environment *sufficient to cause* MS in them |
| $P(E \vert G, E_T)$ | Probability that the event $(E)$ occurs during $(E_T)$ for a *proband* randomly-selected from $(G)$ |
| $u$ | Variable, which represents the level of environmental-exposure as measured by the odds that the event $(E)$ occurs during *any* $(E_T)$ |
| $a$ | Level of environmental-exposure during *some* specific $(E_T)$ – i.e., when: $(u = a)$ |
| $h(u), k(u)$ | Unknown (and unspecified) hazard functions for *susceptible-men* – $h(u)$; and for *susceptible-women* – $k(u)$ |
| $H(a), K(a)$ | Cumulative hazard functions for *susceptible-men* – $H(a)$; and for *susceptible-women* – $K(a)$<br>– Defined as the definite integrals of these unknown and unspecified hazard functions from an *exposure-level* of: $(u = 0)$ to an *exposure-level* of: $(u = a)$. |
| $R > 0$ | Value of the proportionality-factor (if the hazards are proportional)<br>– i.e., $k(u) = R * h(u)$ |
| $R^{app}$ | The “apparent” value of $R$ – i.e., the value of $R$ for proportional hazards when: $(c = d \leq 1)$ |
| $\lambda_w, \lambda_m$ | Environmental-exposure thresholds for developing MS in *susceptible women* $(\lambda_w)$ and *susceptible men* $(\lambda_m)$ – *see Text* |
| $\lambda$ | Difference in the environmental-exposure threshold between *susceptible women* and *susceptible men*: i.e., $\lambda = \lambda_w - \lambda_m$ |

## Methods

### 1. Genetic-Susceptibility

A population (*Z*), consists of *N* individuals. The “*genetically-susceptible*” (*G*)-subset includes *everyone* who has *any* non-zero chance of developing MS under *some* environmental-conditions. Each of the (*m* ≤ *N*) individuals in the (*G*)-subset (*i* = 1, 2,. .., *m*) has a unique-genotype [*G*_*i*_]. The probability of the event that a *proband*, randomly-selected from (*Z*), is a (*G*)-subset member is: [*P*(*G*) = *m*⁄*N*]. Membership in (*G*) is assumed independent of the environmental-conditions during *any* specific *Time-Period* (*E*_*T*_) – *see legend Table-2; considering the definition of* (*E*_*T*_).

**Table 2.**
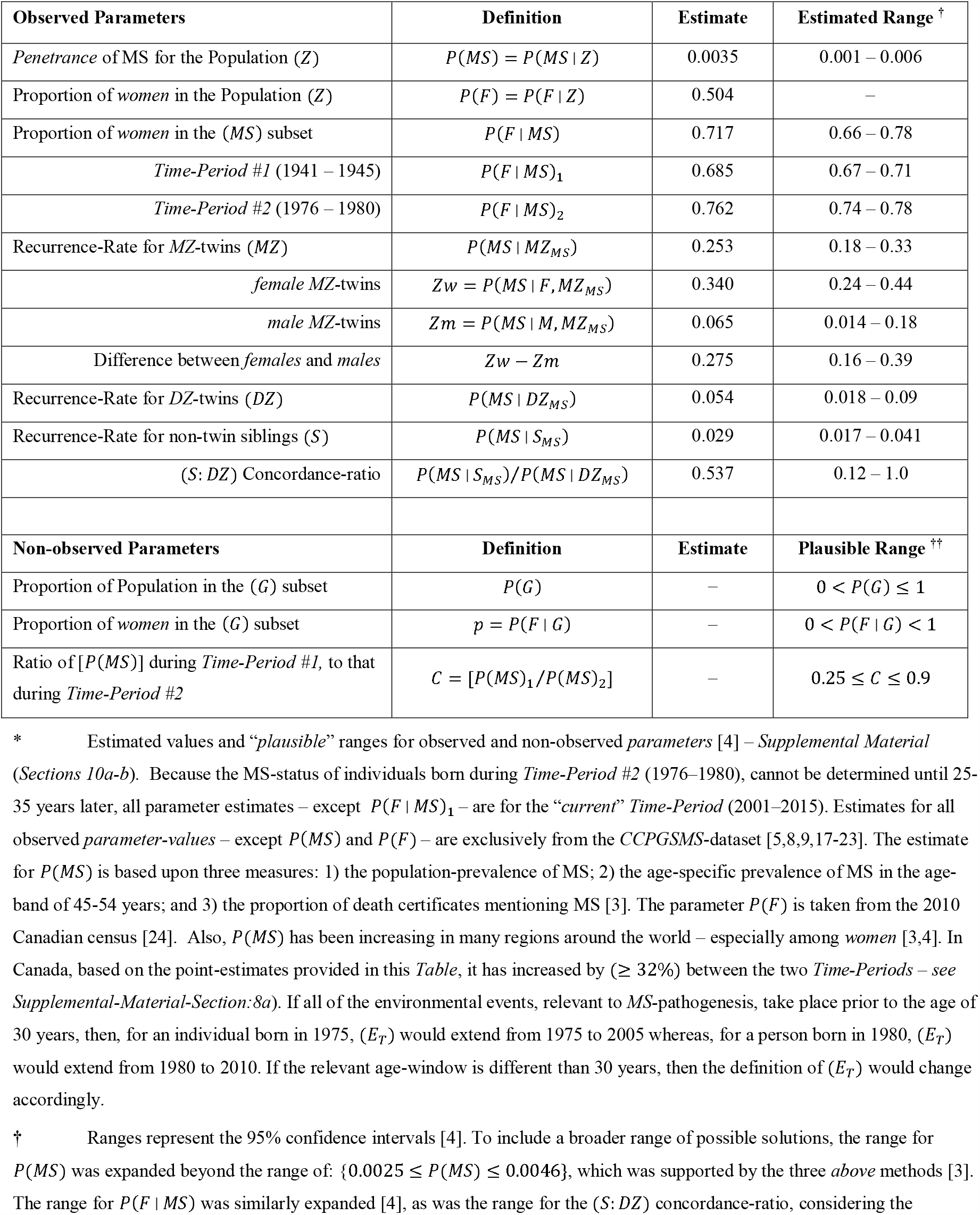
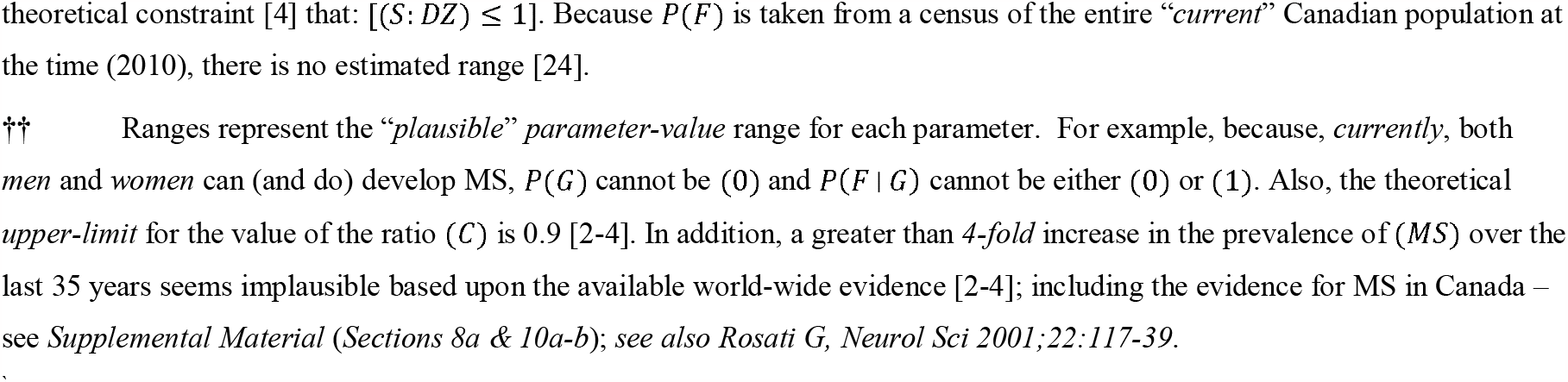
Parameter-values. Point Estimates and Plausible Ranges *

| Observed Parameters | Definition | Estimate | Estimated Range <sup>†</sup> |
| --- | --- | --- | --- |
| Penetrance of MS for the Population ($Z$) | $P(MS) = P(MS \vert Z)$ | 0.0035 | 0.001 – 0.006 |
| Proportion of *women* in the Population ($Z$) | $P(F) = P(F \vert Z)$ | 0.504 | – |
| Proportion of *women* in the ($MS$) subset | $P(F \vert MS)$ | 0.717 | 0.66 – 0.78 |
| *Time-Period #1* (1941 – 1945) | $P(F \vert MS)_1$ | 0.685 | 0.67 – 0.71 |
| *Time-Period #2* (1976 – 1980) | $P(F \vert MS)_2$ | 0.762 | 0.74 – 0.78 |
| Recurrence-Rate for MZ-twins ($MZ$) | $P(MS \vert MZ_{MS})$ | 0.253 | 0.18 – 0.33 |
| *female* MZ-twins | $Zw = P(MS \vert F, MZ_{MS})$ | 0.340 | 0.24 – 0.44 |
| *male* MZ-twins | $Zm = P(MS \vert M, MZ_{MS})$ | 0.065 | 0.014 – 0.18 |
| Difference between *females* and *males* | $Zw - Zm$ | 0.275 | 0.16 – 0.39 |
| Recurrence-Rate for DZ-twins ($DZ$) | $P(MS \vert DZ_{MS})$ | 0.054 | 0.018 – 0.09 |
| Recurrence-Rate for non-twin siblings ($S$) | $P(MS \vert S_{MS})$ | 0.029 | 0.017 – 0.041 |
| ($S:DZ$) Concordance-ratio | $P(MS \vert S_{MS})/P(MS \vert DZ_{MS})$ | 0.537 | 0.12 – 1.0 |
| Non-observed Parameters | Definition | Estimate | Plausible Range <sup>††</sup> |
| Proportion of Population in the ($G$) subset | $P(G)$ | – | $0 < P(G) \leq 1$ |
| Proportion of *women* in the ($G$) subset | $p = P(F \vert G)$ | – | $0 < P(F \vert G) < 1$ |
| Ratio of [$P(MS)$] during *Time-Period #1*, to that during *Time-Period #2* | $C = [P(MS)_1/P(MS)_2]$ | – | $0.25 \leq C \leq 0.9$ |

\* Estimated values and “*plausible*” ranges for observed and non-observed *parameters* [4] – *Supplemental Material* (Sections 10a-b). Because the MS-status of individuals born during *Time-Period #2* (1976–1980), cannot be determined until 25–35 years later, all parameter estimates – except $P(F | MS)_1$ – are for the “*current*” *Time-Period* (2001–2015). Estimates for all observed *parameter-values* – except $P(MS)$ and $P(F)$ – are exclusively from the *CCPGSMS*-dataset [5,8,9,17-23]. The estimate for $P(MS)$ is based upon three measures: 1) the population-prevalence of MS; 2) the age-specific prevalence of MS in the age-band of 45–54 years; and 3) the proportion of death certificates mentioning MS [3]. The parameter $P(F)$ is taken from the 2010 Canadian census [24]. Also, $P(MS)$ has been increasing in many regions around the world – especially among *women* [3,4]. In Canada, based on the point-estimates provided in this *Table*, it has increased by ( $\geq 32\%$ ) between the two *Time-Periods* – see *Supplemental-Material-Section:8a*). If all of the environmental events, relevant to *MS*-pathogenesis, take place prior to the age of 30 years, then, for an individual born in 1975, ( $E_T$ ) would extend from 1975 to 2005 whereas, for a person born in 1980, ( $E_T$ ) would extend from 1980 to 2010. If the relevant age-window is different than 30 years, then the definition of ( $E_T$ ) would change accordingly.
† Ranges represent the 95% confidence intervals [4]. To include a broader range of possible solutions, the range for $P(MS)$ was expanded beyond the range of: $\{0.0025 \leq P(MS) \leq 0.0046\}$ , which was supported by the three *above* methods [3]. The range for $P(F | MS)$ was similarly expanded [4], as was the range for the ( $S:DZ$ ) concordance-ratio, considering the
theoretical constraint [4] that: $[(S:DZ) \leq 1]$ . Because $P(F)$ is taken from a census of the entire “*current*” Canadian population at the time (2010), there is no estimated range [24].
†† Ranges represent the “*plausible*” *parameter-value* range for each parameter. For example, because, *currently*, both *men* and *women* can (and do) develop MS, $P(G)$ cannot be (0) and $P(F \mid G)$ cannot be either (0) or (1). Also, the theoretical *upper-limit* for the value of the ratio ( $C$ ) is 0.9 [2-4]. In addition, a greater than *4-fold* increase in the prevalence of (*MS*) over the last 35 years seems implausible based upon the available world-wide evidence [2-4]; including the evidence for MS in Canada – see *Supplemental Material (Sections 8a & 10a-b)*; see also Rosati G, *Neurol Sci* 2001;22:117-39.

The (*MS*)-subset includes *everyone* who either has, or will subsequently develop, MS. The probability of the event that a *proband*, randomly-selected from (*Z*), is both an (*MS*)-subset member and whose relevant-exposures occurred during (*E*_*T*_), is called the *MS-penetrance* for the population (*Z*) during (*E*_*T*_), or *P*(*MS*|*E*_*T*_). Also, during (*E*_*T*_), the probability of the event that a *proband*, randomly-selected from (*G*), is an (*MS*)-subset member, is called the *MS-penetrance* for the (*G*)-subset during (*E*_*T*_), or *P*(*MS*|*G, E*_*T*_). Both *penetrance-values* depend upon the environmental-conditions during (*E*_*T*_). Also defined are the subsets of *susceptible-women* (*F, G*) and *susceptible-men* (*M, G*). Their *MS-penetrance-values*, during (*E*_*T*_), are:

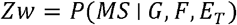

and: 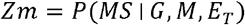.

These *MS-penetrance-values*, (*Zw*) and (*Zm*), are also called the “*failure-probabilities*” for *susceptible-women* and *susceptible-men* during (*E*_*T*_). During *any* (*E*_*T*_), because the proportion of *women* in (*G*) is independent of environmental-conditions, the (*F:M*)-*sex-ratio* always reflects the ratio of these two *failure-probabilities* (*see Supplemental-Material-Section:5d*).

### 2. Environmental-Susceptibility

For each (*G*)-subset member, a family of exposures is defined that includes *every* set of environmental-exposures, each of which is “*sufficient*”, by itself, to *cause* MS in that person. Moreover, for *any* susceptible-individual to develop MS, that person must experience at least one of the “*sufficient-exposure-sets*” within *their* family. Individuals sharing the same family of *sufficient-exposures* – although possibly requiring different “*critical-exposure-intensities*” [4] – belong to the same “*exposure-group*”.

Certain environments may be *sufficient* to *cause* MS in *anyone* but are so improbable (e.g., intentional inoculation of someone with myelin proteins or other agents) that, effectively, they never occur spontaneously. Nevertheless, even individuals who can *only* develop MS under such extreme (or unlikely) conditions, are still (*G*)-subset members – i.e., they *can* develop MS under *som*e environmental-conditions.

The probability of the event (*E*) – i.e., that a randomly-selected member of (*G*), during (*E*_*T*_), experiences an environment *sufficient* to *cause* MS in them – is represented as: *P*(*E*|*G, E*_*T*_). A mathematical definition for the (*E*)-event is provided in the *Supplemental-Material-Section:1a*.

Each set of *sufficient-exposures* is *completely* undefined and agnostic regarding: 1) how many environmental-exposures are involved; 2) when, during life, and in what order, these exposures need to occur; 3) the intensity and duration of the required exposures; 4) what these exposures are; 5) whether *any* of these exposures needs to interact with *any* genetic-factors; and 6) whether certain exposures need to be present or absent. The *only* requirement is that each *exposure-set*, taken together, is *sufficient*, by itself, to *cause* MS in a specific susceptible-individual or in susceptible-individuals belonging to the same *exposure-group*.

## 3. MZ-Twins, DZ-Twins, and Siblings

The term (*MZ*) represents the event that a *proband*, randomly-selected from (*Z*), is an (*MZ*)-subset member or, equivalently, is an *MZ-*twin. This *proband*’s twin is called their “*co-twin*”. The probability that the *proband* belongs to the (*MS, MZ*)-subset and their *co-twin* belongs to (*MZ*) is the same as the probability that the *proband* belongs to (*MZ*) and their *co-twin* belongs to (*MS, MZ*). Therefore, for clarity, (*MS, MZ*) indicates this subset (or event) for the *proband*, whereas (*MZ*_*MS*_) indicates the same subset (or event) for their *co-twin*. The analogous subsets (or events) for *DZ “co-twins*” (*DZ*_*MS*_) and non-twin “*co-siblings*” (*S*_*MS*_) are defined similarly (*Table-1*).

Consequently, *P*(*MS*|*MZ*_*MS*_) represents the life-time probability that a randomly-selected *proband* belongs to (*MS, MZ*), given that their *co-twin* belongs to (*MZ*_*MS*_) – a probability estimated by the *proband-wise* (or *case-wise*) *MZ*-twin-concordance-rate [25].

The term *P*(*MS*|*IG*_*MS*_) represents this concordance-rate – i.e., *P*(*MS*|*MZ*_*MS*_) – adjusted because *MZ*-twins, in addition to sharing “identical” genotypes (*IG*), also share intrauterine and, probably, other environments. This adjustment – made by multiplying the *proband-wise MZ*-twin-concordance-rate by the (*S*: *DZ*) concordance-ratio [4] – isolates the genetic-contribution to the observed *MZ-*twin concordance-rates (*see Supplemental-Material-Section:2a*).

## 4. Estimating P(G)

If the population (*Z*) and the (*G*)-subset are identical, then, during *any* (*E*_*T*_), the *MS-penetrance* of (*Z*) and (*G*) are also identical. Consequently, the ratio of these two *penetrance-values* [4] estimates *P*(*G*) such that:

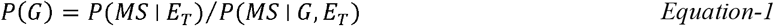

If this ratio is (1), then *everyone* in the population can develop MS under *<u>some</u>* environmental-conditions. However, if the *MS-penetrance* of (*G*) exceeds that of (*Z*), then this ratio is less than (1), indicating that only *some* members of (*Z*) have *any* possibility of developing MS, regardless of *any* exposure they either have had or could have had. Even if the “*exposure-probability*”, *P*(*E|G, E*_*T*_), never reaches 100% under *any* realistic conditions, if (*Z*) and (*G*) are the same, then this ratio is (1) during *every* (*E*_*T*_). Moreover, in *any* circumstance where: [*p* = *P*(*F*|*G*) ≠ *P*(*F*)], it must be that: (*P*(*G*) < 1).

## 5. Data-Analysis

*Cross-sectional-Models use* data from the “*current*” (*E*_*T*_) – *Table-2. Longitudinal-Models* use data regarding changes in MS-epidemiology, which have occurred over the last century [3,4,23] – *see also Supplemental-Material; Figure-S1. Cross-sectional-Models* make the two common assumptions that: 1) *MZ-twinning* is independent of genotype and: 2) *MS-penetrance* is independent of (*MZ*)-subset membership (*Supplemental-Material-Section:4a*). *Longitudinal-Models* make neither assumption. Initially, “*plausible-value-ranges*” are defined for both “observed” and “non-observed” *parameter-values* (*Table-2*). Subsequently, a “substitution-analysis” determined those *parameter-value-combinations* (solutions) that fall within the “*plausible-value-ranges*” for each *parameter*. For each *Model*, (∼10^11^) possible *parameter-value-combinations* were systematically-interrogated.

C*urrently, MS-penetrance* for *female-probands*, whose *co-twin* belongs to (*MZ*_*MS*_), is (5.7)-fold greater than *MS-penetrance* for comparable *male-probands* (*Table-2*). Moreover, *currently*, both the (*F:M*)-*sex-ratio* and *P*(*MS*) are increasing [2-4,23]. Under such circumstances, almost certainly, the *current MS-penetrance* in *susceptible-women* exceeds that in *susceptible-men* (*see Supplemental-Material-Sections:3a&7g*). Therefore, it is assumed that, *currently*:

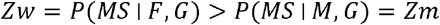

No assumptions are made regarding the circumstances of other *Time-Periods*

## 6. Cross-sectional Models

For notational simplicity, *parameter-abbreviations* are used: *MS-penetrance* for the *i*^*th*^ susceptible individual is: {*x*_*i*_ = *P*(*MS|G*_*i*_)} ; the set (*X*) consists of *MS-penetrance* values for all susceptible-individuals – i.e., (*X*) = (*x*_1_, *x*_2_,*…, x*_*m*_) ; the variance of (*X*) is: 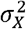; *MS-penetrance* for the (*G*)-subset is: *x* = *P*(*MS|G*) ; the adjusted *MZ-*twin concordance-rate is: *x*’ = *P*(*MS*|*IG*_*MS*_).

During *any* (*E*_*T*_), the *MS-penetrance* for (*Z*) is *P*(*MS*). As demonstrated in the *Supplemental-Material-Section:4a*, during *any* (*E*_*T*_), the *MS-penetrance* for (*G*) is:

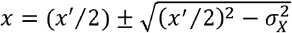

Consequently, during *any* (*E*_*T*_), the probability *P*(*G*) is estimated by the ratio of these *penetrance-values* (*Methods #4*).

## 7. Longitudinal Models

### General Considerations

Using standard survival-analysis methods [26], the exposure (*u*) is defined as the *odds* that the event (E) occurs for a randomly-selected member of the (*G*)-subset during *any Time-Period* (*see Supplemental-Material-Sections:5a-c*). Hazard-functions in *men, h*(*u*), and *women, k*(*u*), are defined in the standard manner [26] and, if these unknown hazard-functions are proportional, a proportionality-factor (*R* > 0) is defined such that:

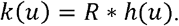

The *exposure-level* (*u* = *a*), during *some Time-Period*, is then converted into “*cumulative-hazard-functions*”, *H*(*a*) and *K*(*a*), which represent definite integrals of these unspecified hazard-functions from an *exposure-level* of: (*u* = 0) to an *exposure-level* of: (*u* = *a*).

*{NB: Cumulative-hazard measures exposure, not failure. Failure is the event that the proband develops MS. The mapping of* (*u* = *a*) *to H*(*a*) *or K(a), if proportional, is “one-to-one and onto” [4]. Therefore, both exposure-measures are equivalent. However, the failure-probabilities (Z*_*w*_ *and Z*_*m*_*) are exponentially related to cumulative-hazard and, therefore, are mathematically-tractable, despite the underlying hazard-functions being unspecified. Moreover, <u>any</u> two points on <u>any</u> exponential curve defines the entire curve*.*}*

Unlike true-survival, for MS, the *failure-probability* may not approach 100% as the *exposure-probability* approaches unity (*see Supplemental-Material-Sections:5b-e*). Moreover, the limiting-value for this *failure-probability* in *susceptible-men* (***c***) and *susceptible-women* (***d***) may not be the same. Also, (***c***) and (***d***) are constants, estimated from the *parameter-values* of (*Zw*), (*Zm*), *P*(*MS*), and the (*F:M*)-*sex-ratio* “observed” during *any* two *Time-Periods*.

The *exposure-level* at which MS becomes possible (i.e., the threshold) must be zero for *susceptible-women* or *susceptible-men* or both. The difference between the threshold in *women* (*λ*_*w*_) and that in *men* (*λ*_*m*_) is defined as:

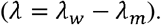

And, therefore: if: *(λ*_*w*_ > *λ*_*m*_); then (*λ*) is positive and (*λ*_*m*_ = 0)

if: *(λ*_*w*_ < *λ*_*m*_); then *(λ*) is negative and *(λ*_*w*_ = 0)

if: *(λ*_*w*_ = *λ*_*m*_); then: *(λ* = *λ*_*w*_ *=λ*_*m*_ = 0)

As demonstrated in the *Supplemental-Material-Section:7a*, if hazards are proportional and if: [*H*(*a*) ≥ λ)], the *cumulative-hazards* in *men* and *women* are related such that:

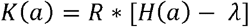

Moreover, any *causal-chain* leading to disease can *only* include genetic-factors, environmental-events, or both (including *any* interactions). Therefore, if *any* member of (*G*) experiences an exposure *sufficient* to *cause* MS in them, and if, in this circumstance, this person’s probability of developing (*MS*) is less than (100%); then their outcome, in part, must be due to a “*truly*” random mechanism. Consequently, if randomness plays *no role* in MS-pathogenesis, then: (***c*** = ***d*** = 1) – *see Discussion*.

Also, regardless of proportionality, *any* disparity between *women* and *men* in their likelihood of developing MS, during *any Time-Period*, must be due to a difference between (***c***) and (***d***), between *susceptible-men* and *women* in the likelihood of their experiencing a *sufficient-exposure*, or between both (*Supplemental-Material-Section:5d*). Therefore, assuming that: (***c*** = ***d*** ≤ 1), also assumes that any difference between *men* and *women* in their *failure-probability* is due, exclusively, to a difference in their likelihood of experiencing a *sufficient-exposure*.

### Non-proportional Hazard

If hazards in *women* and *men* are not proportional, the “*plausible-parameter-value”* ranges still limit possible solutions. However, any difference that these *values* take during different *Time-Pe*r*iods* could be attributed, both potentially and plausibly, to the different environmental-circumstances of different times and places.

### Proportional Hazard

The “*apparent*” value of (*R*), or (*R*^*app*^), is defined as *the* value of (*R*) when: (***c*** =***d*** ≤1). As demonstrated in *Supplemental-Material-Sections:7c&7g*, for proportional-hazards with proportionality-factor (*R*), three conditions must hold:

1. if: *R*≤ 1 ; or, if: *R*< *R*^*app*^ ; or, if: *λ*≤0 ; then: ***c***< ***d***
2. if: ***c*** =***d***≤1 ; then, both: *R*> 1 and: *λ*> 0
3. if: R> 1 ; then: *λ* > 0

*Condition #1* excludes *any* possibility that: (***c*** =***d ≤*** 1) – *see Figures-1&2 and Results*.

**Figure 1.**
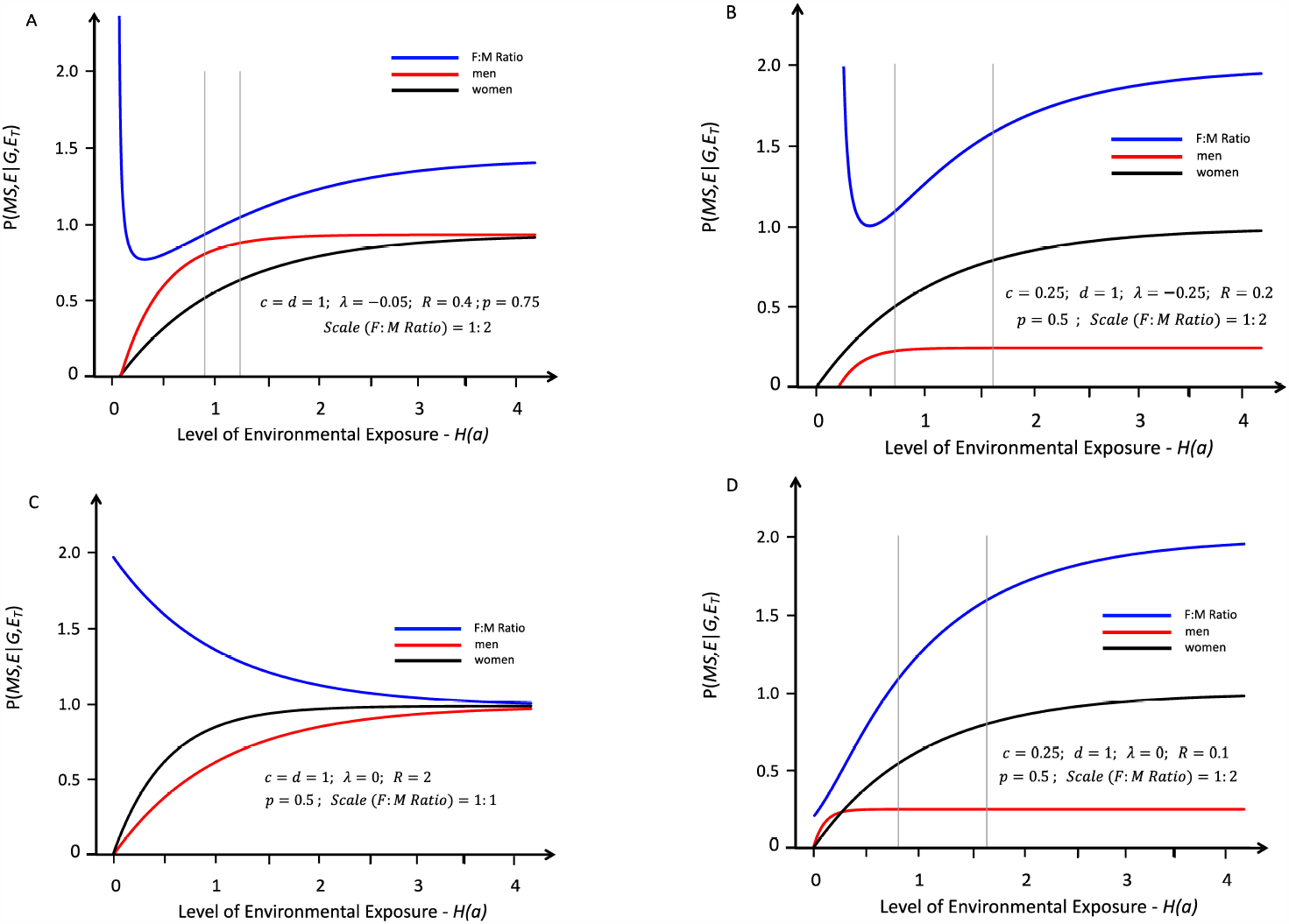
Response-curves representing the likelihood of developing MS in genetically-susceptible *women* (black lines) and *men* (red lines) with an increasing probability of a *sufficient* environmental exposure – *see Methods #2 & Supplemental-Material-Section:1a*. The curves depicted in *Panels A* and *B* are proportional, with a proportionality-factor (*R*), although the environmental threshold is greater for *men* than for *women* – i.e., under conditions in which: (*λ* < 0) – *see Text*. The curves depicted in *Panels C* and *D* are “strictly” proportional, meaning that the environmental threshold is the same for both *men* and *women* – i.e., under conditions in which: (*λ* = *λ*_*W*_ = *λ*_*m*_ = 0) – *see Text*. The blue lines represent the change in the (*F:M*)*-sex-ratio* (plotted at various scales; indicated in each *Panel*) with increasing exposure. The thin grey vertical lines represent the portion of the response-curve that covers the change in the (*F:M*)*-sex-ratio* from 2.2 to 3.2 (i.e., the actual change observed in Canada [23] between *Time-Periods #1 & #2*). The grey lines are omitted in *Panel C* because the observed (*F:M*)-*sex-ratio* change is not possible under these conditions. In *Panel A*, although the (*F:M*)-*sex-ratio* change is possible, the condition (*Zw* > *Zm*) is never possible throughout the entire response-curve. Response-curves *A, B*, and *D* reflect conditions in which (*R* < 1); whereas curve *C* reflects conditions in which (*R*> 1). If (*R*= 1), the blue line in *Panel C* would be flat (*see Supplemental-Material-Sections;7c-f*). Response-curves *A* and *C* reflect conditions in which (***c*** =***d***= 1); whereas curves *B* and *D* reflect those conditions in which (***c***< ***d***= 1).

**Figure 2.**
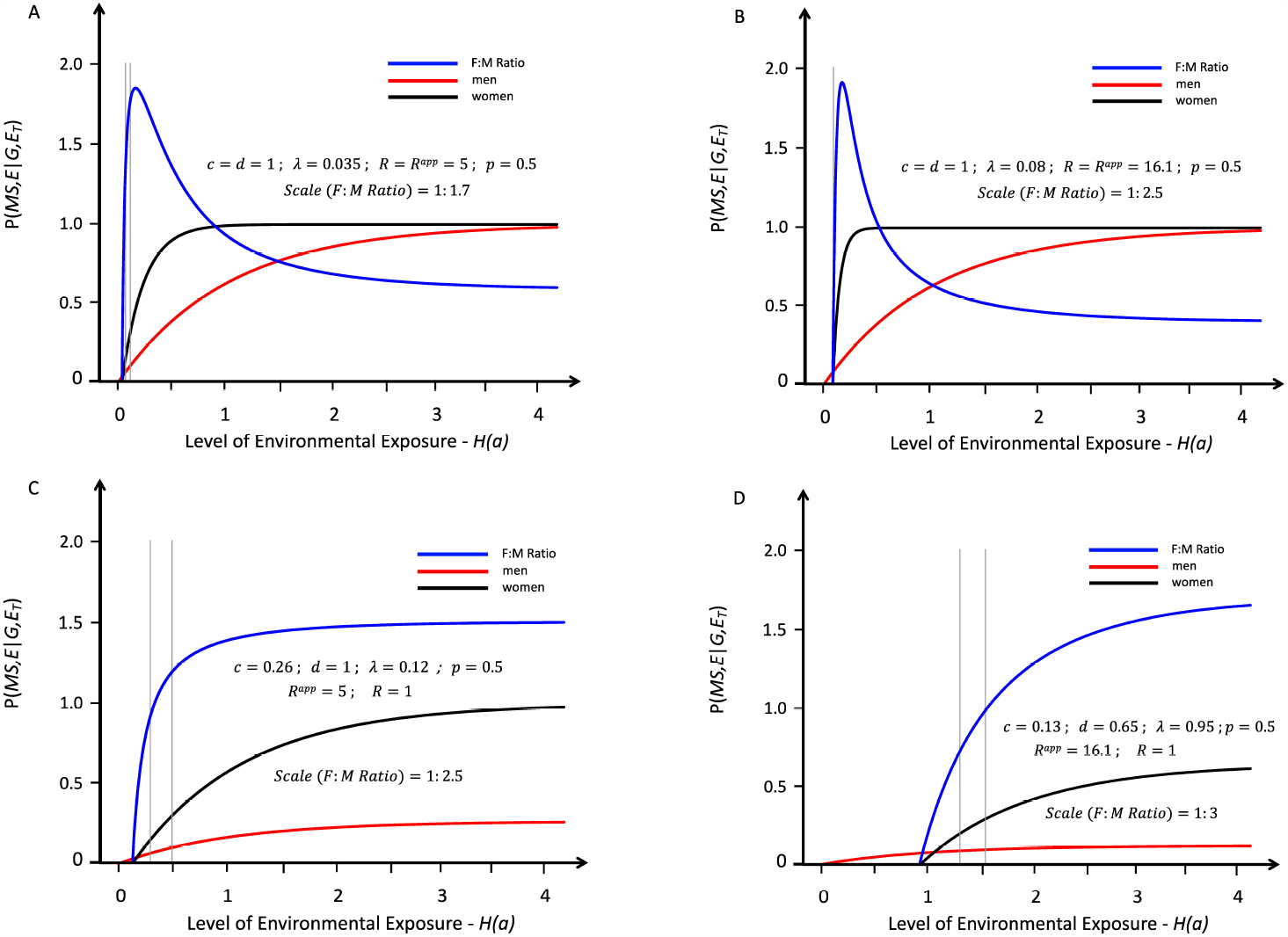
Response-curves for the likelihood of developing MS in genetically *susceptible-women* (black lines) and *men* (red lines) with an increasing probability of a *sufficient* environmental exposure – *see Methods #2 & Supplemental-Material-Section:1a*. The curves depicted are proportional, with a proportionality-factor (*R*), although the environmental threshold is greater for *women* than that it is in *men* – i.e., these are conditions in which: (*λ*> 0). Also, all of these response-curves represent actual solutions. The blue lines represent the change in the (*F:M*)*-sex-ratio* (plotted at various scales; indicated in each *Panel*) with increasing exposure. *Panels A* and *B* are for conditions where: (***c*** =***d*** = 1). The value of (*R*), specific for this condition, is termed (*R*^*app*^). Indeed, for *every* condition in which: (***c***=***d ≤*** 1), both: (*R* = *R*^*app*^) and the response curves for *men* and *women* have the same relationship to each other (*see Supplemental-Material-Sections:7c-f*). By contrast, *Panels C* and *D* represent conditions where: (***c*** <***d ≤*** 1) and, in these circumstances: (*R* < *R*^*app*^). To account for the observed increase in the (*F:M*)*-sex-ratio*, the response-curves in Panels *A* and *B require* that the Canadian observations [23] were made within a very narrow window – i.e., for most of these response-curves, the (*F:M*)*-sex-ratio* is actually decreasing. By contrast, the response-curves in *Panels C* and *D* demonstrate an increasing (*F:M*)*-sex-ratio* for *every* two-point interval of exposure along the entire response-curves for *women* and *men*. The thin grey vertical lines represent the portion of these response-curves (for the depicted solution), which represents the actual change in the (*F:M*)*-sex-ratio* for specific “solutions” between *Time-Periods #1 & #2*.

*Conditions #2&3* (where: *λ*> 0), require that, as the *odds* of a *sufficient-exposure* decreases, there must come a point where only *susceptible-men* can develop MS. This implies that, at (or below) this *sufficient-exposure-level*, (*R* = 0). Consequently, the additional requirement that: (*R* > 1) poses a potential paradox.

There are two obvious ways to avoid this paradox (*see Supplemental-Material-Sections:7d-h*). First, if the hazards are non-proportional, although this creates other problems. For example, *women* and *men* in the same *exposure-group*, necessarily, have proportional-hazards (*Supplemental-Material-Section:7h*). Therefore, if *women* and *men* are never in the same *exposure-group*, each sex *must* develop MS in response to distinct {*E*_*i*_} families, in which case *female-MS* and *male-MS* would represent different diseases.

Second, *Condition #1* is compatible with any (*λ*) so that, if: (*λ*> 0) and (*R* ≤1), then, at every *sufficient-exposure-level* (*u* = *a*), the probability that a *susceptible-man*, randomly-selected, will experience a *sufficient-exposure* is as great, or greater, than this probability for a *susceptible-woman*.

## Results

### 1. Cross-sectional Models

For all *Cross-sectional* analyses [4], the supported-range for *P*(*G*) is:

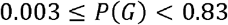

From *Equation-1*, assuming: (*x* ≥*x*^’^⁄2); the supported-range for P(G) is:

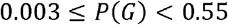

### 2. Longitudinal Models

For either non-proportional or proportional-hazards and, if proportional, any (*R*), the supported-range for *P*(*G*) is:

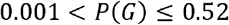

For proportional-hazards and: (***c*** =***d*** = 1), the supported ranges for the threshold-difference between *susceptible-women* and *susceptible-men* (*λ*); for the proportionality-factor (*R* = *R*^*app*^); and the *probability-ratio* for receiving a *sufficient*-exposure *–* i.e., *P*(*E*∣*F, G*)⁄*P*(*E*|*M, G*) *–* are:

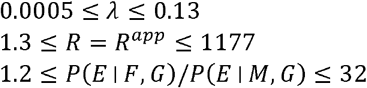

For proportional-hazards and both (*R* = 1) & (***d*** = 1), the supported-ranges for (*λ*) and for the limiting probability of developing MS in *susceptible-men* (*c*) are:

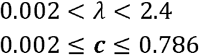

## Discussion

The two principal conclusions from this analysis are: 1) the *penetrance* of (*G*) is greater than that of (*Z*) and, thus, *not everyone* in the population is susceptible to developing MS and: 2) at *maximum exposure-levels*, the limiting probability for developing MS in *susceptible-men* (***c***) is less than that for *susceptible-women* (***d***). These two conclusions, stated explicitly, are:

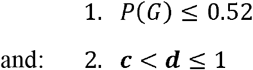

*Conclusion #1* seems inescapable (*see Results*). Indeed, given *any* of the reported *MZ-*twin-concordance-rates, the notion that *MS-penetrance* for (*G*) is the same as that for (*Z*) is untenable (*Table-4; Reference:[3]*). Therefore, a large proportion of the population (*Z*) must be impervious to developing MS, regardless of *any* environmental-events they either have experienced or could have experienced.

Regarding *Conclusion #2*, however, scenarios exist where: (***c*** = ***d*** ≤ 1) might be possible. Principal among these is the possibility of non-proportional-hazards, which requires *female-MS* and *male-MS* to be different diseases (*Methods #7*). However, given the genetic and environmental evidence, this too seems untenable. For example, all but one of the 233 MS-associated loci are *autosomal*, and the single *X-chromosome* risk-variant is present in both sexes [6] – *see also Supplemental-Material-Section:7f*. Moreover, the MS-association with different *HLA-*haplotypes is the same for both sexes – *see Tables-3&4; Reference:[4]*. Family studies also suggest a common genetic-basis for MS in *women* and *men* [2-5,8,22,27,28]. Thus, both twin and non-twin siblings (*male* or *female*) of an *MS-proband* have increased MS-risk, regardless of *proband* sex [5,27,28]. Similarly, both *sons* and *daughters* of conjugal couples have markedly increased MS-risk [8,27]. Also, *male* and *female* full-or half-siblings with an MS-*proband* parent (*mother* or *father*) have increased MS-risk [2,8,22,27]. Each of these observations, supports a similar (if not the same) genetic-basis for MS in both sexes.

Also, contrary to those circumstances *<u>required</u>* whenever the proportionality-factor (*R*) is greater than (1) – i.e., when (*R* > 1) – *women* don’t seem more likely than *men* to experience the MS-associated environmental-events (either known or suspected). Thus, for both sexes, the *month-of-birth* effect is equally evident [2,4,9-11]; the latitude gradient is the same [2,4,12]; the impact of intrauterine/perinatal environments is similar (*Supplemental-Material-Section:2c*); *EBV* infection is equally common [2,4,13,14]; vitamin-D levels are the same [2,4,15,16]; and smoking tobacco is actually less common among *women* [2,4]. Collectively, these observations suggest that, *currently*, each sex experiences the relevant environmental-events in an approximately equivalent manner. Taken together, this genetic and environmental evidence implies that *female-MS* and *male-MS* represent the same underlying disease-process and, therefore, that the hazards are proportional (*Methods #7*).

Also, several lines of evidence indicate that the proportional-hazard condition of: (***c*** = ***d*** ≤ 1) is unlikely. First, the environmental-observations (*described above*) suggest that: (*R*^*app*^ > *R* ≈ 1), which is impossible whenever: (***c*** = ***d*** ≤ 1) – *see Methods #7 & Results*. Second, as in *Figure-1*, whenever (λ ≤ 0) or whenever (*R* ≤ 1), the condition that: (***c*** < ***d***) is established (*Methods #7*). Third, the alternative of: (*R* > 1) & (*λ* > 0) creates a potential paradox (*Methods #7*). Although there are ways to rationalize this potential paradox with: (***c*** = ***d*** ≤ 1), in every case, the conditions required *whenever*: (***c*** < ***d*** ≤ 1) are far less extreme [4]. Finally, the response-curves when: (***c*** = ***d*** ≤ 1) & (*R* > 1) are steeply ascending and present only a very narrow exposure-window to explain the (*F:M*)*-sex-ratio* data [23] – *see Figures-2A&2B*. Moreover, following this narrow-window, the (*F:M*)*-sex-ratio* decreases with increasing exposure. By contrast, the Canadian MS-data documents a steadily-progressive rise in the (*F:M*)*-sex-ratio* over the past century [4,23] – *see also Supplemental-Material; Figure-S1*.

Nevertheless, whenever (***c*** < ***d***), some *susceptible-men* will *never* develop MS, even when a susceptible-genotype co-occurs with a *sufficient-exposure*. Thus, the Canadian MS-data [5,8,9,17-23] seems to indicate that MS-pathogenesis involves a “*truly*” random element. This cannot be attributed to other, unidentified, environmental-factors (e.g., other infections, diseases, nutritional deficiencies, toxic-exposures, etc.) because each set of environmental-exposures is defined to be *sufficient*, by itself, to *cause* MS in a specific susceptible-individual. If other conditions were necessary for this individual to develop MS, then one (or more) of their *sufficient-exposure sets* would include these conditions (*Methods #2*). This also cannot be attributed to the possibility that *some* individuals can *only* develop MS under improbable conditions. Thus, the estimates for (***c***) and (***d***) are based solely upon observable *parameter-values* (*Methods #7*). Finally, this cannot be attributed to mild or asymptomatic-disease because this *disease-type* occurs disproportionately among *women* compared to the *current* (*F:M*)-*sex-ratio* in MS [4,23]. Naturally, invoking “*truly*” random-events in disease-expression requires replication. Nevertheless, *any* finding that: (***c*** < ***d***) indicates that the behavior of some complex physical-systems (e.g., organisms) involve “*truly*” random-mechanisms.

Moreover, considering those circumstances where: (*R* = 1) & (***d*** = 1) and, also, considering a *man*, randomly-selected from the (*M, G*)-subset, who also experiences a *sufficient-environment*, the chance that he *will not* develop MS is (21 − 99%) – *see Results*. Consequently, both the genetic and environmental data, which support the conclusion that: (*R* ≈ 1) – *see above* – also, support the conclusion that it is this “random-element” of disease-pathogenesis, which is primarily responsible for the difference in disease-expression currently-observed between *susceptible-women* and *men*. Importantly, the fact that a process favors disease-development in *women* over *men* does *<u>not</u>* imply that the process *must be* non-random. For example, when flipping a biased-coin compared to a fair-coin – if both are random-processes – the only difference is that, for the biased-coin, the two possible outcomes are not equally likely. In the context of MS-pathogenesis, the characteristics of “*female-ness*” and “*male-ness*” would each simply be envisioned as biasing the coin differently (whatever characteristics are implied by these two terms).

Other authors, modeling immune-system function, also invoke random-events in MS-disease-expression [4]. In these cases, however, randomness is incorporated into their *Models* to reproduce the MS-disease-process more faithfully. However, the fact that including randomness improves a *model’s* performance doesn’t constitute a *test* of whether “*true*” randomness ever occurs. For example, the outcome of a dice-roll may be most accurately *modeled* by treating this outcome as a random-variable with a well-defined probability-distribution. Nevertheless, the question remains whether this probability distribution represents a *complete* description of the process, or whether this distribution is merely a convenience, compensating for our ignorance regarding the initial orientation of the dice and the direction, location, and magnitude of the forces that act on the dice during the roll [4,29,30].

It is hard to imagine that the outcomes of complex-biological-processes such as evolution and immune-function are pre-determined events, especially considering the fact that both processes are so remarkably adaptive to contemporary external-events [4,30]. Nevertheless, *proving* that any macroscopic-process is “*truly*” random is difficult. This requires an experiment (i.e., a *test*), in which the outcome predicted by determinism differs from that predicted by non-determinism.

The Canadian MS-data presents an opportunity to apply just such a *test*. Thus, the widely-held deterministic-hypothesis [4,30] *<u>requires</u>* that: (***c*** = ***d*** = 1). *Any* observation that either: (***c*** < ***d*** = 1) or: (***c*** ≤ ***d*** < 1) indicates that “*true*” randomness is a component of disease-development and undermines the deterministic-view. Therefore, the Canadian MS-data [5,8,9,17-23], which strongly implies that: (c < d), provides empirical evidence in support of the non-deterministic hypothesis. Importantly, this analysis *explicitly includes* all those genetic-factors and environmental-events (including interactions), which are necessary for MS-pathogenesis, regardless of whether these factors, events, and interactions are known, suspected, or as yet unrecognized. Nevertheless, in addition to these necessary prerequisites, “*true*” randomness also seems critical to disease-pathogenesis. Moreover, both sexes seem to have the same underlying-disease. Thus, both seem to have a similar genetic-basis and, also, a similar response to the same environmental disease-determinants (*see above*). These observations suggest both that the hazards are proportional (*Methods #7*) and that (*R* ≈1). If correct, this indicates that it is this “*truly*” random-element in disease-pathogenesis, which is primarily responsible for the currently-observed differences in disease-expression between *susceptible-women* and *susceptible-men*.

## Data Availability

All data is available in the published literature

## Acknowledgements

I am especially indebted to John Petkau, PhD, Professor Emeritus, Department of Statistics, University of British Columbia, Canada, for enormous help with this project. He devoted many hours of his time to critically reviewing early versions of this analysis and contributed immensely both to the clarity and to the logical development of the mathematical and statistical arguments presented herein. I am also indebted to my mentor, Michael J. Aminoff, MD, Professor Emeritus, Department of Neurology, University of California, San Francisco, USA, for his invaluable help with this project. He critically, and thoughtfully, reviewed many drafts of this manuscript and contributed enormously to the logic and clarity of its presentation.

