## Supplementary material for "Pathogenesis of Multiple Sclerosis: Genetic, Environmental, and Random Mechanisms": Supplemental Material.pdf

***Mathematical Development of the Susceptibility Models***

***For Drfinitions of Model Terms – see Section 9; Tables S1a-b***

**1. *Environmental Susceptibility***

### 1. *Environmental Susceptibility*

#### *1a. Defining Environmental Susceptibility* ($E$)

The population $(Z)$ consists of $(N)$ individuals. The "*genetically-susceptible*" subset $(G)$ – i.e., the subset of *everyone* who has *any* non-zero chance of developing MS under *some* environmental circumstances – consists of $(m \leq N)$ individuals $(i = 1,2, \dots, m)$, each having a unique genotype $[G_i]$. Even $MZ$-twins, despite having "*identical*" genotypes $(IG)$, still have subtle genetic differences from one another [4]. For the purpose of this analysis, it is assumed these subtle differences are unrelated to susceptibility [4]. The probability of the event that an individual, randomly selected from $(Z)$, is a member of the $(G_i)$ subset – a subset consisting of a single individual – is: $[P(G_i) = 1/N]$. The *MS-penetrance* for this subset $(x_i)$, during $(E_T)$, is: $[x_i = P(MS \mid G_i, E_T)]$. By the definition of $(G)$, *above*, it *must* be that: $(\forall\, G_i \in (G){:}\;\; x_i > 0)$ under *some* environmental conditions.

The family $\{E_i\}$ includes *every* set of environmental exposures, each of which is "*sufficient*", by itself, to *cause* MS to develop in the *i*^th^ susceptible individual (including *any* necessary interactions between genes and environment). Each "*sufficient*" *exposure-set* within the $\{E_i\}$ family must be distinct (in some way) from each other although, otherwise, there can be any degree of overlap between the exposures that comprise these sets. Moreover, the $\{E_i\}$ family can contain an unlimited number of "*sufficient*" *exposure-sets* although, because: $[\forall\, G_i \in (G){:}\, x_i > 0]$ under *some* environmental conditions, the family cannot be empty. The event $\{E_i\}$ indicates that, at least, one of these "*sufficient*" *exposure-sets* within the $\{E_i\}$ family occurs. Moreover, it is possible that two or more members of $(G)$ may share the same $\{E_i\}$ family of exposures – although perhaps requiring different "*critical exposure intensities*" [4]. If so, such individuals are said to belong to the same "*i-type*" *exposure-group*.

For the *i*^th^ susceptible individual to develop MS, the events $\{E_i\}$ and $(G_i)$ *must* occur jointly – i.e., the individual $(G_i)$ *must* experience one or more of the $\{E_i\}$ environments. This joint occurrence is reflected by the subset $(\{E_i\}, G_i)$ and the occurrence of $(\{E_i\}, G_i)$ represents the event that an individual, selected randomly from $(Z)$ – the *proband* – is both the *i*^th^ susceptible individual and that they experience an $\{E_i\}$ environment "*sufficient*" to cause MS in them. The probability of this event, given that this person is in $(G)$ and given the environmental conditions of $(E_T)$, is represented as $P(\{E_i\}, G_i \mid G, E_T)$. If the event $(G_i)$ occurs without $\{E_i\}$, then whatever exposure does occur, it is *insufficient*, and the *i*^th^ susceptible individual cannot develop MS.

The event $(E)$ is defined to be the union of the disjoint events, which exhibit the pairing of the $(m)$ susceptible individuals with their "*sufficient*" *exposure-sets*, such that:

$$(E) = (\{E_1\}, G_1) \cup (\{E_2\}, G_2) \cup \dots \cup (\{E_m\}, G_m)$$

in which case: $$P(E \mid G, E_T) = \textstyle\sum_{i=1}^{m} P(\{E_i\}, G_i \mid G, E_T)$$

or: $$P(E \mid G, E_T) = \textstyle\sum_{i=1}^{m} P(G_i \mid G, E_T) * P(\{E_i\} \mid G_i, G, E_T)$$

Because genotype is independent of the environmental conditions of $(E_T)$:

$$\forall\, G_i \in (G){:}\;\; P(G_i \mid G, E_T) = P(G_i \mid G) = \frac{P(G_i)}{P(G)} = \frac{(1/N)}{(m/N)} = 1/m$$

so that: $$P(E \mid G, E_T) = (1/m) * \textstyle\sum_{i=1}^{m} P(\{E_i\} \mid G_i, G, E_T)$$

In this way, the term $P\big(E \,\big|\, G, E_T\big)$ represents the probability of the event that an individual, selected randomly from the $(G)$-subset, experiences an environmental exposure during $(E_T)$, which is "*sufficient*" to *cause* MS in them. Furthermore, by definition, the event $(E)$ can only occur in circumstances where the event $(G)$ also occurs. Therefore: $P(E, G) = P(E)$

### 2. *Adjusting the MZ-twin Concordance for the Shared Environment of Twins*

#### *2a. Adjustment for the Shared Environment of MZ-twins* – $\boldsymbol{P(MS \,|\, IG_{MS})}$

By definition, anyone with MS <u>*must*</u> belong to $(G)$ and <u>*must*</u> have experienced the event $(E)$. Therefore:

$$P(MS) = P(MS, G) = P(MS, E) = P(MS, E, G)$$

so that: $$P(MS \,|\, MZ_{MS}) = P(MS, E, G \,|\, MZ_{MS}) = \sum_{i=1}^{m} P\big(MS, \{E_i\}, G_i \,\big|\, MZ_{MS}\big)$$

where: $\forall(i)$: $(i = 1,2, \dots, m)$:

$$P(MS, \{E_i\}, G_i \,|\, MZ_{MS}) = P(MS \,|\, \{E_i\}, G_i, MZ_{MS}) * P(\{E_i\} \,|\, G_i, MZ_{MS}) * P(G_i \,|\, MZ_{MS})$$

In this manner, the probability that the *proband* is a member of the $(MS, \{E_i\}, G_i)$ subset, given the fact that their *co-twin* is a member of the $(MZ_{MS})$ subset – i.e., $P(MS, \{E_i\}, G_i \,|\, MZ_{MS})$ – can be deconstructed and re-expressed as the product of three component probabilities – 1) the probability that MS develops in an *MZ-proband* $(G_i)$ who experiences a "*sufficient*" exposure $\{E_i\}$; 2) the probability that this *MZ-proband* experiences an $\{E_i\}$ exposure, which is "*sufficient*" to *cause* MS in them; and 3) the probability that this *MZ-proband* is a member of the $(G_i)$-subset – where each probability is conditioned on fact that the *proband* has an *MZ co-twin*, who is a member of the $(MZ_{MS})$ subset within $(Z)$ – *see Main Text*.

For *probands* who are members of $(G)$, but who are otherwise unspecified, the analogous probabilities can be written:

$$P(MS \,|\, G) = P(MS, E, G \,|\, G) = \sum_{i=1}^{m} P\big(MS, \{E_i\}, G_i \,\big|\, G\big)$$

where: $\forall(i)$: $(i = 1,2, \dots, m)$:

$$P(MS, \{E_i\}, G_i \,|\, G) = P(MS \,|\, \{E_i\}, G_i) * P(\{E_i\} \,|\, G_i) * P(G_i \,|\, G)$$

Therefore, to determine the necessary adjustment, the impact of *MZ*-twins sharing environments needs to be removed while, at the same time, leaving the genetic impact of being *MZ*-twins unchanged. To this end, one can define the term $(IG_{MS})$ such that:

$$P(MS, \{E_i\}, G_i \,|\, IG_{MS}) = P(MS \,|\, \{E_i\}, G_i, IG_{MS}) * P(\{E_i\} \,|\, G_i, IG_{MS}) * P(G_i \,|\, IG_{MS})$$

where: $P(\{E_i\} \,|\, G_i, IG_{MS}) = P(\{E_i\} \,|\, G_i)$

and: $P(G_i \,|\, IG_{MS}) = P(G_i \,|\, MZ_{MS})$

Moreover, the conditioning events $(\{E_i\}, G_i)$ and $(\{E_i\}, G_i, MZ_{MS})$ both represent the same underlying event for the *proband* – i.e., the event that the *i*^*th*^ susceptible individual (the *proband*) experiences an environment "*sufficient*" to cause MS in them. In this circumstance, therefore:

$$P(MS \mid \{E_i\}, G_i, MZ_{MS}) = P(MS \mid \{E_i\}, G_i, IG_{MS}) = P(MS \mid \{E_i\}, G_i)$$

Incorporating these equivalences, into the *above* definition of $(IG_{MS})$, yields:

$$P(MS, G_i \mid IG_{MS}) = P(MS, \{E_i\}, G_i \mid IG_{MS}) = P(MS \mid \{E_i\}, G_i) * P(\{E_i\} \mid G_i) * P(G_i \mid MZ_{MS})$$

or: $$P(MS, G_i \mid IG_{MS}) = P(MS \mid G_i) * P(G_i \mid MZ_{MS}) = (x_i) * P(G_i \mid MZ_{MS})$$

In this manner, the *above* definition for $(IG_{MS})$ can be re-expressed such that:

$$\forall\ G_i \in (G): \quad \frac{P(MS, G_i \mid IG_{MS})}{P(G_i \mid IG_{MS})} = P(MS \mid G_i, IG_{MS}) = P(MS \mid G_i) = x_i$$

and $$\forall\ G_i \in (G): \quad P(G_i \mid IG_{MS}) = P(G_i \mid MZ_{MS})$$

And, thus, the appropriate "*adjusted*" probability, $P(MS \mid IG_{MS})$, can be expressed as:

$$P(MS \mid IG_{MS}) = \sum_{i=1}^{m} P(MS, G_i \mid IG_{MS}) = \sum_{i=1}^{m} P(G_i \mid MZ_{MS}) * (x_i)$$

This adjustment, effectively, represents a thought-experiment, in which susceptible *MZ*-twins are separated at conception, and where the *proband* twin is expected to experience the same environmental exposure as would any $(G)$-subset member, given the environmental conditions of $(E_T)$.

*{NB: This definition represents the intended meaning of the "adjusted" proband-wise (or case-wise) recurrence rate [25] for MZ-twins – i.e.,* $P(MS \mid IG_{MS})$*. The appropriate adjustment can be made such that:*

$$s_a = P(MS \mid DZ_{MS})/P(MS \mid S_{MS})$$

*and:* $$P(MS \mid IG_{MS}) = P(MS \mid MZ_{MS})/s_a$$

*as demonstrated in the Supplementary Material of Reference #4.}*

#### *2b. Adjustment for the Susceptible Women and Men Considered Together*

***Assertion:*** $P(MS \mid IG_{MS}) = 0.136$

***Proof:*** The following point-estimates (*Table 2; Main Text; see also Section 10b; Table S2; below*) from the Canadian twin-study [5] will be used:

$$P(MS \mid MZ_{MS}) = 0.253$$
$$P(MS \mid DZ_{MS}) = 0.054$$
$$P(MS \mid S_{MS}) = 0.029$$

From the *Supplementary Material* (*Reference #4*), one can estimate the *point-value* of $\{P(MS \mid IG_{MS})\}$ as:

$$s_a = P(MS \mid DZ_{MS})/P(MS \mid S_{MS}) = 0.054/0.029 = 1.86$$

and: $$P(MS \mid IG_{MS}) = P(MS \mid MZ_{MS})/s_a = 0.253/1.86 = 0.136$$

*2c. Adjustments for Susceptible Women and Men Considered Separately*

***Assertions:*** $P(MS \mid F, IG_{MS}) = P(MS \mid F, MZ_{MS})/1.95$

$P(MS \mid M, IG_{MS}) = P(MS \mid M, MZ_{MS})/1.63$

***Proof:*** Two parameters ($s_{aw} \geq 1$) and ($s_{am} \geq 1$) are defined such that:

$$P(MS \mid F, IG_{MS}) = P(MS \mid F, MZ_{MS})/s_{aw}$$

$$P(MS \mid M, IG_{MS}) = P(MS \mid M, MZ_{MS})/s_{am}$$

From the point-estimates of the Canadian epidemiological data [5,8,17-23] – see *Section 10b; below* – and from *Assertion 4A* (*Section 4a; below*), therefore:

$$P(F \mid MS) = P(F \mid IG_{MS}) = 0.717$$

$$P(F \mid MS, MZ_{MS}) = P(F \mid MS, IG_{MS}) = 0.917$$

$$P(MS \mid F, MZ_{MS}) = 0.340$$

$$P(MS \mid M, MZ_{MS}) = 0.065$$

The term, $P(MS, F \mid IG_{MS})$, can be deconstructed in two different ways:

$$P(MS, F \mid IG_{MS}) = P(F \mid IG_{MS}) * P(MS \mid F, IG_{MS}) = (0.717 * 0.340)/s_{aw}$$

and: $$P(MS, F \mid IG_{MS}) = P(MS \mid IG_{MS}) * P(F \mid MS, IG_{MS}) = (0.136 * 0.917)$$

Combining these two equations leads to:

$$s_{aw} = (0.717 * 0.340)/(0.136 * 0.917) = 1.95$$

Similarly: $$P(MS, M \mid IG_{MS}) = P(M \mid IG_{MS}) * P(MS \mid M, IG_{MS}) = (0.283 * 0.065)/s_{am}$$

and: $$P(MS, M \mid IG_{MS}) = P(MS \mid IG_{MS}) * P(M \mid MS, IG_{MS}) = (0.136 * 0.083)$$

leading to: $$s_{am} = (0.283 * 0.065)/(0.136 * 0.083) = 1.63$$

Thus, the point estimate for the impact of *MZ-twins* sharing their intrauterine and *some* of their childhood environments on the likelihood that the *proband* twin is a member of ($MS$), given the fact that their *co-twin* a member of the ($MZ_{MS}$), is very similar for both *women* and *men*.

### 3. *Enrichment of Women among MS-Patients and Concordant MZ- Twins*

#### *3a. Enrichment of More Penetrant Genotypes*

If the *MS-penetrance* for susceptible *women* exceeds that in *men* (i.e., $Zw > Zm$) then, from the *Supplementary Material* (*Reference #4*), from the definition of $(IG_{MS})$ – *see Section 2a; above* – and from *Assertion 4A* (*below*), *women* can be described as being "*enriched*" such that:

$$P(F|G, MS, MZ_{MS}) = P(F|G, MS, IG_{MS}) > P(F|G, MS) > P(F|G)$$

The terms $(G_{i1})$ and $(G_{i2})$ represent the events that *any* pair of *probands*, randomly selected from $(G)$, belong, respectively, to the $(G_{i1})$ and $(G_{i2})$ subsets – each subset consisting of a single individual. The probability of each of these events – *see Section 1a; above* – is:

$$P(G_{i1}|G) = P(G_{i2}|G) = 1/m$$

The *MS-penetrance values* of these two subsets are designated, respectively, as:

$$x_{i1} = P(MS|G, G_{i1}) \quad \text{and:} \quad x_{i2} = P(MS|G, G_{i2})$$

Moreover, these two subsets can be suitably defined such that: $(x_{i1} \geq x_{i2})$.

For notational simplicity, the following probability terms [including the definition of $(IG_{MS})$ – *Section 2a* (*above*) – and from *Assertion 4A; below*] are defined such that:

$$p = P(F|G) \ ; \ x = P(MS|G) \ ; \ x' = P(MS|IG_{MS}) \ ; \ Zw = z_w \ ; \ Zm = z_m$$

$$z_w = P(MS|F, G) \ ; \ z'_w = P(MS|F, G, IG_{MS}) \ ; \ z_m = P(MS|M, G) \ ; \ z'_m = P(MS|M, G, IG_{MS})$$

$$x'_{i1} = P(MS|G_{i1}, IG_{MS}) = x_{i1} \ ; \ x'_{i2} = P(MS|G_{i2}, IG_{MS}) = x_{i2}$$

***Assertion:*** Almost certainly**:** $Zw = P(MS|F, G) > P(MS|M, G) = Zm$

***Development:*** With respect to the subsets $(G_{i1})$ and $(G_{i2})$, therefore:

$$P(G_{i1}|G, MS) = \frac{P(G_{i1}, G, MS)}{P(G, MS)} = \frac{P(G_{i1}|G) * P(MS|G, G_{i1})}{P(MS|G)} = (G_{i1}|G) * (x_{i1}/x)$$

and: $$P(G_{i2}|G, MS) = \frac{P(G_{i2}, G, MS)}{P(G, MS)} = \frac{P(G_{i2}|G) * P(MS|G, G_{i2})}{P(MS|G)} = (G_{i2}|G) * (x_{i2}/x)$$

Therefore: $\forall\, G_{i1} \ \& \ G_{i2} \in (G)$: $P(G_{i1}|G, MS) \geq P(G_{i2}|G, MS)$

Also: $$P(G_{i1}|G, MS, IG_{MS}) = \frac{P(G_{i1}, G, MS, IG_{MS})}{P(G, MS, IG_{MS})} = \frac{P(G_{i1}|G, IG_{MS}) * P(MS|G, G_{i1}, IG_{MS})}{P(MS|G, IG_{MS})}$$

From the definition of $(IG_{MS})$ – *Section 2a* (*above*) – and *Assertion 4A* (*below*), therefore

$$P(G_{i1}|G, MS, IG_{MS}) = P(G_{i1}|G, MS) * (x_{i1}/x')$$

and similarly: $$P(G_{i2}|G, MS, IG_{MS}) = P(G_{i2}|G, MS) * (x_{i2}/x')$$

Thus: $\forall\, G_{i1} \,\&\, G_{i2} \in (G)$: $P(G_{i1}|G, MS, IG_{MS}) \geq P(G_{i2}|G, MS, IG_{MS})$

Therefore, within the $(MS, G)$ subset, genotypes are "*sorted*" in the sense that the most prevalent genotypes are also the most penetrant for *every pair-wise* comparison. Similarly, within the $(MS, G, IG_{MS})$ subset, this "*sorting*" is even more extreme for *every pair-wise* comparison and, therefore, there is a continuing "*enrichment*" of more penetrant genotypes such that:

$$\forall\ G_{i1}\ \&\ G_{i2} \in (G):\ \ 1 = \frac{P(G_{i1} \mid G)}{P(G_{i2} \mid G)} \leq \frac{P(G_{i1} \mid G,MS)}{P(G_{i2} \mid G,MS)} \leq \frac{P(G_{i1} \mid G,MS,IG_{MS})}{P(G_{i2} \mid G,MS,IG_{MS})}$$

Moreover, using the terminology of *Section 7h* (*below*) to specify members of the $(G)$ subset, the $(mp)$ members of the $[(F, G) = (G_w)]$ subset are designated such that: $(d = 1,2, \dots, mp)$, each with a unique genotype $(G_{dw})$, an *MS-penetrance value* of $(z_{dw})$, and a variance for the set of these *penetrance values* of $(\sigma_w^2)$. Analogously, the $[m(1-p)]$ members of the $[(M, G) = (G_m)]$ subset can be designated such that: $[d = 1,2, \dots, m(1-p)]$, each with a unique genotype $(G_{dm})$, an *MS-penetrance* value of $(z_{dm})$, and a variance for the set of these *penetrance values* of $(\sigma_m^2)$. In this case:

$$z_w = P(MS \mid F, G) = \sum_{dw=1}^{mp} P(MS, G_{dw} \mid F, G) = \sum_{dw=1}^{mp} P(G_{dw} \mid F, G) * (z_{dw}) = E(z_{dw})$$

and also: $z_m = P(MS \mid M, G) = E(z_{dm})$

Similarly: $P(MS \mid F, G, IG_{MS}) = \sum_{dw=1}^{mp} P(MS, G_{dw} \mid F, G, IG_{MS}) = \sum_{dw=1}^{mp} P(G_{dw} \mid F, G, IG_{MS}) * (z_{dw})$

where: $P(G_{dw} \mid F, G, IG_{MS}) = P(G_{dw} \mid F, G) * (z_{dw})/P(MS \mid F, G)$

so that: $z_w' = E[(z_{dw})^2]/z_w = z_w + \sigma_w^2/z_w$

Following the logic of the *Assertion 4B* proof (*below*), therefore: $z_w = (z_w'/2) \pm \sqrt{(z_w'/2)^2 - \sigma_w^2}$

And also: $z_m' = E[(z_{dm})^2]/z_m = z_m + \sigma_m^2/z_m$ so that: $z_m = (z_m'/2) \pm \sqrt{(z_m'/2)^2 - \sigma_m^2}$

Both $P(MS)$ and the (*F:M*) *sex ratio* are currently increasing [1-4,23] – *see also Sections 8a & 10a-b* (*below*). Therefore, also, *currently*, $(Zw)$ must be increasing at a faster rate than $(Zm)$ – *see Section 7g* (*below*). Moreover, the MS data from Canada [5] – *see Section 10b* (*below*) – indicate that *currently*:

$$P(MS \mid F, MZ_{MS})_2 = (5.7) * P(MS \mid M, MZ_{MS})_2 \quad \& \quad P(F \mid MS)_2 = 0.762$$

Therefore, unless $[P(F \mid G) \geq P(F \mid MS)_2]$ – or, equivalently, unless: $[p/(1-p)] \geq$ *the current* (*F:M*) *sex ratio* (*see Equation S5j; below*) – and unless susceptible *men* and *women* have markedly different and non-unimodal *variance-distributions* for their *MS-penetrance values* [2-4], then, currently, it <u>*must*</u> be the case that:

$$z_w = Zw = P(MS \mid F, G) > P(MS \mid M, G) = Zm = z_m$$

Moreover, if susceptible *men* and *women* can both be members of every "*i-type*" *exposure-group* (*see Sections 7g-h; below*), it would be very hard to rationalize such an extreme difference in *variance-distributions*. Consequently, we assume that this relationship pertains during the "*current*" *Time Period.*

*{NB: Because the observations regarding $(Zw)$ and $(Zm)$, presented in the Main Text (Table 2), only relate to the "current" Time Period, the circumstances of other Time Periods cannot be determined.}*

4. ***Cross-sectional Model***

***4a. Model Development***

For notational simplicity, the following probability terms are defined:

$$p = P(F \mid MS)\ ; \quad x_i = P(MS \mid G_i);\ x_i'' = P(MS \mid G_i, MZ_{MS})\ ;\ x = P(MS \mid G)\ ;\ \text{and: } x' = P(MS \mid IG_{MS})$$

***Assertions:*** ***4A.*** $\forall\ G_i \in (G)$: $P(G_i, MS \mid MZ) = P(G_i, MS)$

$\forall\ G_i \in (G)$: $P(G_i \mid IG_{MS}) = P(G_i \mid MZ_{MS}) = P(G_i \mid MS)$

$P(IG_{MS}) = P(MZ_{MS}) = P(MS)$

$P(F \mid IG_{MS}) = P(F \mid MZ_{MS}) = P(F \mid MS)$

$P(F \mid MS, IG_{MS}) = P(F \mid MS, MZ_{MS})$

***4B.*** $x = (x'/2) \pm \sqrt{(x'/2)^2 - \sigma_X^2}$

***4C.*** $0 \leq \sigma_X^2 \leq (x'/2)^2$

$\sigma_X^2 = x(x' - x)$

***Definitions and Assumptions:*** The subset $(G)$ is defined (*see Main Text & Section 1a*) and, as noted:

$$\forall\ G_i \in (G):\ \ x_i = P(MS \mid G_i)$$

Thus, $(x_i)$ represents the *MS-penetrance* for the *i^th^* susceptible individual whose exposure occurs during any specific *Time Period* and it is unique to the *i^th^* individual. The set $(X)$ is defined to include the *penetrance-value* for each of the $(m)$ members of the $(G)$ subset – i.e., $(X) = (x_1, x_2, \ldots, x_m)$ – and its variance is defined to be $(\sigma_X^2)$. Finally, each of the $(k)$ individuals in the population $(k = 1, 2, \ldots, N)$ has a unique genotype $\{G_k\}$ – including *MZ*-twins who, despite sharing "*identical*" genotypes, still have subtle genetic differences from one another [4].

A random variable $(x_G)$ can be defined to represent any of the $\{x_i\}$ elements within the set $(X)$ and from this, and from *Section 1a* (*above*), the following terms can be defined:

$P(G) = m/N$

$\forall\ G_i \in (G)$: $P(G_i \mid G) = 1/m$

$E(x_G) = \sum_{i=1}^{m} (x_i) * (1/m) = P(MS \mid G) = x$ (*Equation S4a*)

$E(x_G^2) = \sum_{i=1}^{m} (x_i^2) * (1/m) = x^2 + \sigma_X^2$ (*Equation S4b*)

$x' = P(MS \mid IG_{MS}) = P(MS, G \mid G, IG_{MS}) = \sum_{i=1}^{m} P(MS, G_i \mid G, IG_{MS})$ (*Equation S4c*)

These *Equations*, and those derived *below*, describe relationships for the subset $(G)$. In a similar manner, analogous relationships can be established and derived for the subsets $(F, G)$ and $(M, G)$ – *see Section 3a; above – see also Supplemental Material; Reference #4.*

Two assumptions are made:

*Assumption #1*

*MZ*-twinning is generally thought to be non-hereditary [4]. If so, then every person (i.e., genotype) in the population $(Z)$ has the same chance, *a priori*, of having an *MZ*-twin (i.e., *MZ*-status is independent of genotype). In this circumstance, during any *Time Period*, it will be the case that:

$$\forall\, G_k \in (Z):\; P(MZ \,|\, G_k) = P(MZ)$$

and, thus: $$\forall\, G_i \in (G):\; P(MZ \,|\, G_i) = P(MZ)$$

Even if *MZ*-twinning were thought to be hereditary in some circumstances [4], but where those genetic factors, which relate to *MZ*-twinning, are independent of MS-susceptibility, then the same conclusion would follow. Either this, or the *above* condition, are assumed to pertain.

*Assumption #2*

The *MS-penetrance* for *any proband MZ*-twin (whose *co-twin* is of unknown status) is assumed to be independent of *MZ*-status. Thus, this *penetrance-value* for any genotype is presumed to be the same regardless of whether that genotype occurs with or without having an *MZ co-twin*. This assumption is equivalent to assuming that experiencing any particular environment together with an *MZ co-twin* has the same impact as experiencing that environment alone. Alternatively, it is presumed that the mere fact of having an *MZ co-twin* does not alter the environment in such a way that the development of MS becomes more or less likely in both the *proband* and the *co-twin*. Specifically, it is assumed, for any *Time Period*, that:

$$\forall\, G_i \in (G):\; P(MS \,|\, G_i, MZ) = P(MS \,|\, G_i)$$

***Proof of Assertion 4A:***

From *Assumption #1*, it follows that:

$$\forall\, G_k \in (Z):\quad P(G_k, MZ) = P(G_k) * P(MZ \,|\, G_k) = P(G_k) * P(MZ)$$

and therefore: $$\forall\, G_k \in (Z):\; P\big(G_k \,|\, MZ\big) = P(G_k, MZ)/P(MZ) = P(G_k)$$

Consequently, also: $$\forall\, G_i \in (G):\; P\big(G_i \,|\, MZ\big) = P(G_i)$$

From this conclusion, from the definitions of $(MZ_{MS})$ – *see Main Text* – and from *Assumption #2*, it follows that, during any *Time Period*:

$$\forall\ G_i \in (G):\ P(G_i, MZ_{MS}) = P\big(MS, G_i \,\big|\, MZ\big) = P\big(G_i \,\big|\, MZ\big) * P\big(MS \,\big|\, G_i, MZ\big)$$

$$= P(G_i) * P(MS \,|\, G_i) = P(MS, G_i)$$

and: $$P(MZ_{MS}) = P(MS \,|\, MZ) = \textstyle\sum_{i=1}^{m} P(\,G_i \,|\, MZ) * P(MS \,|\, G_i, MZ)$$

$$= \textstyle\sum_{i=1}^{m} P(G_i) * P(MS \,|\, G_i) = P(MS)$$

From the definition of $(IG_{MS})$ – *see Section 2a* (*above*) – and from these two equivalences, one can conclude that, during any *Time Period*:

$$\forall\ G_i \in (G):\quad P\big(G_i \,\big|\, IG_{MS}\big) = P\big(G_i \,\big|\, MZ_{MS}\big) = \frac{P(G_i, MZ_{MS})}{P(MZ_{MS})} = \frac{P(G_i, MS)}{P(MS)} = P\big(G_i \,\big|\, MS\big)$$

Consequently: $\forall\ G_i \in (G)$:

$$P(G_i, IG_{MS}) = P(IG_{MS}) * P\big(G_i \,\big|\, MS\big) = P(MZ_{MS}) * P\big(G_i \,\big|\, MS\big) = P(G_i, MZ_{MS})$$

From this result, together with the conclusion from (*above*), therefore, both:

$$P(IG_{MS}) = P(MZ_{MS}) = P(MS) \qquad \text{and also:} \qquad P(G_i, IG_{MS}) = P(G_i, MZ_{MS})$$

Moreover, from the definition of $(IG_{MS})$ – *see Section 2a; above* – it follows that:

$$P(MS \,|\, MZ_{MS}) = (s_a) * P(MS \,|\, IG_{MS})$$

Therefore: $$P(MS \,|\, MZ_{MS}) = \textstyle\sum_{i=1}^{m}\ (s_a) * P(MS, G_i \,|\, IG_{MS}) = \sum_{i=1}^{m} P(G_i \,|\, IG_{MS}) * [\,(s_a) * (x_i)]$$

and also: $$P(MS \,|\, MZ_{MS}) = \textstyle\sum_{i=1}^{m}\ P(MS, G_i \,|\, MZ_{MS}) = \sum_{i=1}^{m} P(G_i \,|\, MZ_{MS}) * (x_i'')$$

Consequently, from *above*: $\forall\ G_i \in (G)$:

$$P(G_i \,|\, IG_{MS}) * [(s_a) * (x_i)] = P(G_i \,|\, MZ_{MS}) * [(s_a) * (x_i)] = P(G_i \,|\, MZ_{MS}) * (x_i'')$$

so that: $$\forall\ G_i \in (G):\ \ (s_a) * (x_i) = (x_i'') \quad \& \quad (s_a) * P(MS, G_i \,|\, IG_{MS}) = P(MS, G_i \,|\, MZ_{MS})$$

Therefore: $$P(G_i \,|\, MS, IG_{MS}) = P(G_i \,|\, MS, MZ_{MS})$$

Using the terminology of *Section 7h* (*below*) to designate the *women* of $(G)$, it follows that each of these $(mp)$ *women* $(d = 1,2, \ldots, mp)$, has a unique genotype $(G_{dw})$ and, therefore:

$$P(F \,|\, IG_{MS}) = \textstyle\sum_{d=1}^{mp} P(G_{dw} \,|\, IG_{MS}) = \sum_{d=1}^{mp} P(G_{dw} \,|\, MS) = P(F \,|\, MS)$$

and: $$P(F \,|\, MS, IG_{MS}) = \textstyle\sum_{d=1}^{mp} P(G_{dw} \,|\, MS, IG_{MS}) = \sum_{d=1}^{mp} P(G_{dw} \,|\, \text{MS}, MZ_{MS}) = P(F \,|\, \text{MS}, MZ_{MS})$$

similarly: $$P(M \,|\, IG_{MS}) = P(M \,|\, MS) \qquad \text{and:} \qquad P(M \,|\, MS, IG_{MS}) = P(M \,|\, MS, MZ_{MS})$$

***Proof of Assertion 4B:***

From the definitions of $(G)$ & $(IG_{MS})$ – *see Main Text & Section 2a* (*above*) – it follows that:

$$P(MS \mid G_i, IG_{MS}) = P(MS \mid G_i, G, IG_{MS}) = P(MS \mid G_i, G) = P(MS \mid G_i) = x_i$$

Therefore, during any *Time Period*, the probability $P(MS, G_i \mid G, IG_{MS})$ can be re-expressed as:

1. $P(MS, G_i \mid G, IG_{MS}) = P(G_i \mid G, IG_{MS}) * P(MS \mid G_i, G, IG_{MS})$

$$= P(G_i \mid G, IG_{MS}) * (x_i)$$

From *Assertion 4A* and from the definitions of $(G)$ & $(IG_{MS})$ – *see Main Text & Sections 1a & 2a* (*above*) – the term $P(G_i \mid G, IG_{MS})$ can be re-expressed as:

2. $P(G_i \mid G, IG_{MS}) = P(G_i \mid G, MS) = P(G_i, G, MS)/P(MS, G)$

$$= P(MS \mid G_i, G) * P(G_i, G)/P(MS, G)$$

$$= (x_i) * P(G_i \mid G)/P(MS \mid G) = (x_i) * (1/m)/x$$

Combining *1 & 2 (above*) yields:

$$P(MS, G_i \mid G, IG_{MS}) = (x_i)^2 * (1/m)/x$$

However, from *Equations S4b-c*, it is the case that:

$$x' = P(MS, G \mid G, IG_{MS}) = \sum_{i=1}^{m} P(MS, G_i \mid G, IG_{MS})$$

where: $\sum_{i=1}^{m} P(MS, G_i \mid G, IG_{MS}) = \sum_{i=1}^{m} (x_i^2) * (1/m)/x = E(x_G^2)/x$

Therefore, from *Equation S4b*, it follows that:

$$x' = (x^2 + \sigma_X^2)/x = x + \sigma_X^2/x \qquad \text{(\textit{Equation S4d})}$$

Rearrangement of *Equation S4d*, yields a standard-form quadratic *Equation* in ($x$) such that:

$$x^2 - (x')x + \sigma_X^2 = 0$$

which, in turn, can be solved to yield:

$$x = (x'/2) \pm \sqrt{(x'/2)^2 - \sigma_X^2} \qquad \text{(\textit{Equation S4e})}$$

***Proof of Assertion 4C:***

*Equation S4e* has real solutions only for the range of:

$$0 \leq \sigma_X^2 \leq (x'/2)^2 \qquad (\textit{Equation S4f})$$

Notably, the maximum variance ($\sigma^2$) for *any* distribution [*Reference: see footnote #1; below*] on the closed interval $[a, b]$ is:

$$\sigma^2 = [(b - a)/2]^2$$

Consequently, regardless of any *Assumptions* (*see above*), the *variance-range* indicated by *Equation S4f* represents the maximum possible *variance-range* for <u>any</u> distribution on the closed interval of: $[0, x']$.

Also, rearrangement of *Equation S4d* yields:

$$\sigma_X^2 = x(x' - x) \qquad (\textit{Equation S4g})$$

##### *4b. Quadratic Equations for Penetrance in Women and Men*

For notational simplicity, the following probability terms are defined:

$$x = P(MS \mid G)\ ; \quad x' = P(MS \mid G, IG_{MS}) = P(MS \mid IG_{MS})\ ;$$

$$Zw = z_w = P(MS \mid F, G)\ ; \quad z'_w = P(MS \mid F, G, IG_{MS}) = P(MS \mid F, IG_{MS})\ ;$$

$$Zm = z_m = P(MS \mid M, G)\ ; \quad z'_m = P(MS \mid M, G, IG_{MS}) = P(MS \mid M, IG_{MS})\ ;$$

$p = P(F \mid G)$ ; and the two ratios: $r = z'_w/z_w$ and: $s = z'_m/z_m$

***Assertions:*** 1. $$Zw = z_w = \frac{x+\sqrt{x^2- \{1+(r/s)(1-p)/p)\}\{x^2-xx'(1-p)/s}\}}{p+(r/s)(1-p)}$$

2. $$Zm = z_m = \frac{x-\sqrt{x^2- \{1+(s/r)(p/(1-p))\}\{x^2-xx'\,p/r}\}}{(1-p)+(s/r)p}$$

***Proof:*** $$P(MS \mid G) = P(MS, F \mid G) + P(MS, M \mid G)$$

$$= P(F \mid G) * (P(MS \mid F, G) + P(M \mid G) * (P(MS \mid M, G)$$

or: $x = p(z_w) + (1 - p)(z_m)$

with re-arrangement, this becomes:

$$z_m = [x - p(z_w)]/(1 - p) \qquad (\textit{Equation S4h})$$

Also: $$x' = P(MS \mid G, IG_{MS}) = P(MS, F \mid G, IG_{MS}) + P(MS, M \mid G, IG_{MS})$$

---

*#1 Jacobson HI. The maximum variance of restricted unimodal distributions. Ann Math Stat. 1969;40:1746–52.*

Therefore, from *Assertion 4A* (*above*):

$$P(MS,F\,|\,G,IG_{MS}) = P(F\,|\,G,IG_{MS}) * (z'_w) = P(F\,|\,G,MS) * (z'_w)$$

and: $P(MS,M\,|\,G,IG_{MS}) = P(M\,|\,G,IG_{MS}) * (z'_m) = P(M\,|\,G,MS) * (z'_m)$

where: $P(F\,|\,G,MS) = P(F,MS\,|\,G)/P(MS\,|\,G) = p(z_w)/x$

and, similarly: $P(M\,|\,G,MS) = (1-p)(z_m)/x$

so that: $xx' = p(z_w)(z'_w) + (1-p)(z_m)(z'_m) = pr * (z_w)^2 + (1-p)s * (z_m)^2$

or: $(z_m)^2 = [xx' - pr(z_w)^2]/[(1-p)s]$ (*Equation S4i*)

Therefore, there are two simultaneous *Equations* for $(z_m)^2$ – i.e., *Equations S4h and S4i, above*. Using these two estimates to eliminate the $(z_m)$ parameter, yields:

$$[\{x - p(z_w)\}/(1-p)]^2 = (z_m)^2 = [xx' - pr(z_w)^2]/[(1-p)s]$$

or: $\{x - p(z_w)\}^2 = \{xx' - pr(z_w)^2\}(1-p)/s = \{xx'(1-p)/s\} - (r/s)p(1-p)(z_w)^2$

and: $x^2 - 2xp(z_w) + p^2(z_w)^2 - xx'(1-p)/s + (r/s)p(1-p)(z_w)^2 = 0$

This last *Equation* can be rearranged to yield a standard-form quadratic *Equation* in $(z_w)$ such that:

$\{p^2 + (r/s)p(1-p)\}(z_w)^2 - \{2xp\}(z_w) + \{x^2 - xx'(1-p)/s\} = 0$ (*Equation S4j*)

Because: $(z'_w \gg z'_m)$ and because both $P(MS)$ and the (*F:M*) *sex ratio* are "*currently*" known to be increasing [3,4,23] – *see also Sections 8a & 10a-b* (*below*) – it is assumed, during the *current Time Period,* that: $(z_w > z_m)$ – *see Section 3a* (*above*). Therefore, *Equation S4j* is solved for $(z_w)$ as:

$Zw = z_w = \frac{x + \sqrt{x^2 - \{1 + (r/s)(1-p)/p)\}\{x^2 - xx'(1-p)/s\}}}{p + (r/s)(1-p)}$ (*Equation S4k*)

*Equation S4h* (*above*) can then be solved for $(z_m)$. Alternatively, the *above* arguments can be reframed to eliminate $(z_w)$ instead of $(z_m)$, and the resulting quadratic *Equation* can be solved for $(z_m)$ as:

$Zm = z_m = \frac{x - \sqrt{x^2 - \{1 + (s/r)(p/(1-p)\}\{x^2 - xx'\,p/r\}}}{(1-p) + (s/r)p}$ (*Equation S4l*)

***5. Longitudinal Model*:**

***5a. Model Development***

Following standard survival analysis methods [26], the cumulative survival $\{S(u)\}$ and failure $\{F(u)\}$ functions where: $F(u) = 1 - S(u)$ can be defined separately for susceptible *men* $\{S_m(u) \text{ and } F_m(u)\}$ and for susceptible *women* $\{S_w(u) \text{ and } F_w(u)\}$. Also, the (unknown and unspecified) hazard functions for developing MS at different environmental *exposure-levels* $(u)$ – i.e., $h(u)$ and $k(u)$ – can be defined for susceptible *men* and susceptible *women*, respectively. These hazard functions for *women* and *men* may be proportional to each other and, if they are proportional, we can then define a hazard proportionality factor $(R > 0)$ such that: $[k(u) = R * h(u)]$. Furthermore, from *Section 1a* (*above*), the term, $P(E \,|\, G, E_T)$, represents the probability of the event that a *proband*, randomly selected from $(G)$, and who has their relevant exposures during $(E_T)$, experiences an environmental exposure "*sufficient*" to *cause* MS in them. The *exposure-level* $(u)$ is then defined as the odds that this event occurs such that:

$$u = \frac{P(E \,|\, G, E_T)}{[1 - P(E \,|\, G, E_T)]}$$

The cumulative hazard function (for *men*), $H(a)$, is defined as the definite integral of the hazard function, $h(u)$, from an *exposure-level* of $(u = 0)$ to an *exposure-level* of $(u = a)$ such that:

$$H(a) = \int_0^a h(u)du$$

Similarly, the cumulative hazard function (for *women*), $K(a)$, is defined as the definite integral of the hazard function, $k(u)$, from an *exposure-level* of $(u = 0)$ to an *exposure-level* of $(u = a)$ such that:

$$K(a) = \int_0^a k(u)du$$

If these hazards are proportional, then:

$$K(a) = \int_0^a R * h(u)du = R * H(a)$$

For *men*, using the common definition of the hazard function [26] that:

$$h(u) = f_m(u)/S_m(u)$$

together with the fact that, by definition:

$$f_m(u) = d[F_m(u)]/du = -d[S_m(u)]/du$$

a standard derivation from survival analysis methods [26] demonstrates that, for *men*, because:

$$h(u)du = -d[S_m(u)]/S_m(u)$$

Therefore, the cumulative hazard function $[H(a)]$ can be re-expressed such that:

$$H(a) = -\int_0^a d[S_m(u)]/S_m(u) = \ln[S_m(0)] - \ln[S_m(a)]$$

where: $H(0) = \ln[S_m(0)] - \ln[S_m(0)] = 0$

Exposure is here being measured as the odds, during $(E_T)$, that a $(G)$-subset member receives an environmental exposure "*sufficient*" to *cause* MS in them. By definition, when: $[P(E \mid G, E_T) = 0]$, no member of $(G)$ can develop MS [i.e., $S_m(0) = 1$], in which case: $\{\ln[S_m(0)] = \ln(1) = 0\}$. Thus:

$$S_m(a) = e^{-H(a)}$$

This standard derivation from survival methods [26], therefore, demonstrates that the survival function is exponentially related to the integral of the underlying hazard function – i.e., the cumulative hazard function. Consequently, the failure function for susceptible *men* can be stated such that:

$$F_m(a) = 1 - S_m(a) = 1 - e^{-H(a)}$$

#### *5b. Environmental Exposure Levels during Different Time Periods*

*{NB: In this and the Sections that follow, observations made during the two Time Periods are distinguished by the use of subscripts (1) and (2). For example, $P(MS)_1$ refers to $P(MS)$ during the 1st Time Period whereas $P(MS)_2$ refers to $P(MS)$ during the 2nd Time Period. Also, it is important to note that cumulative hazard is being used as a measure of exposure, not failure – see Main Text & Reference #4.}*

The environmental *exposure-level* for susceptible *men* during the *1st Time Period* is defined as $[H(a_1)]$. In turn, the *failure-probability* for a susceptible *man* is defined as: $[F_m(a) = Zm]$, which represents the life-time probability of the event that a susceptible *man*, randomly selected from $(G)$, and who has their relevant exposures during $(E_T)$, develops MS. Moreover, if the constant $(\boldsymbol{c})$ is defined as the maximum possible *failure-probability* for susceptible *men*, then:

$$F_m(a) = Zm = P(MS \mid M, G, E_T) = P(MS, E \mid M, G, E_T)$$

and: $\boldsymbol{c} = \lim_{a \to \infty}(Zm) = P(MS \mid M, G, E) \leq 1$

In this circumstance, this *failure-probability* during the *1st Time Period* $(Zm_1)$, can be stated as:

$$F_m(a_1) = Zm_1 = P(MS, E \mid M, G)_1 = \boldsymbol{c} * [1 - e^{-H(a_1)}] \qquad (\textit{Equation S5a})$$

If the *exposure-level* for susceptible *men* during the *2nd Time Period* is defined as $[H(a_2)]$, then, because $(Zm)$ is *currently* increasing with time [3,4], the difference in the *exposure-level* for *men* between the *1st and 2nd Time Periods* can be represented by the parameter $(q_m)$ such that:

$$H(a_2) - H(a_1) = q_m > 0$$

In this case, the *failure-probability* during the *2nd Time Period* $(Zm_2)$, can be stated as:

$$F_m(a_2) = Zm_2 = P(MS, E \mid M, G)_2 = \boldsymbol{c} * [1 - e^{-\{H(a_1)+q_m\}}] \qquad (Equation\ S5b)$$

*Equations S5a & S5b* can be rearranged to yield:

$$1 - Zm_1/\boldsymbol{c} = e^{-H(a_1)}$$

and: $$1 - Zm_2/\boldsymbol{c} = e^{-\{H(a_1)+q_m\}} \qquad (Equation\ S5c)$$

Dividing the 1st of these two *Equations* by the 2nd yields:

$$(1 - Zm_1/\boldsymbol{c})/(1 - Zm_2/\boldsymbol{c}) = e^{q_m} \qquad (Equation\ S5d)$$

or: $$q_m = \ln(1 - Zm_1/\boldsymbol{c}) - \ln(1 - Zm_2/\boldsymbol{c}) \qquad (Equation\ S5e)$$

This unit $(q_m)$ is arbitrary but, nonetheless, depends upon the actual (but unknown) change in the environmental *exposure-level*, which has taken place between the two *Time Periods*. From *Equations S5d–e*, the estimated magnitude of this *exposure-level* change depends upon the value of $(\boldsymbol{c})$, which can range over the interval of: $(1 \geq \boldsymbol{c} > Zm_2)$. The ratio on the *LHS* of *Equation S5d* (*above*) is always greater than unity because $(Zm)$ increases with increasing exposure. Moreover, it increases monotonically as $(\boldsymbol{c})$ varies throughout its range – being at a minimum when: $(\boldsymbol{c} = 1)$ and approaching infinity as: $(\boldsymbol{c} \rightarrow Zm_2)$.

Consequently, the term $(q_m^{min})$ is defined to be the *"minimum" exposure-level* change that is possible for *men* between these two *Time Periods*. In this case, this *minimum exposure-level* change will occur when:

$$\boldsymbol{c} = P(MS \mid M, E, G) = 1$$

Therefore, from *Equation S5e*: $q_m^{min} = \ln(1 - Zm_1) - \ln(1 - Zm_2)$

Nevertheless, this *minimum exposure-level* change $(q_m^{min})$ may not accurately reflect the actual (but unknown) change in the *exposure-level*, which has taken place between the two *Time Periods*. Therefore, the term $(q_m)$ is called the *"actual" exposure-level* change for susceptible *men*. This may well be different from the "*minimum*" possible *exposure-level* change so that:

$$q_m \geq q_m^{min}$$

In a directly analogous manner, the term $[F_w(a) = Zw]$ is defined to be the *failure-probability* for susceptible *women* during any *Time Period* and the constant $(\boldsymbol{d})$ is defined to be the maximum possible *failure-probability* for susceptible *women* such that:

$$F_w(a) = Zw = P(MS \mid F, G, E_T) = P(MS, E \mid F, G, E_T)$$

and: $$\boldsymbol{d} = \lim_{a \to \infty}(Zw) = P(MS \mid M, F, E) \leq 1$$

Similar to *Equations S5a-b* (*above*), because $(Zw)$ is also increasing with time [3,4], the *failure-probability* in susceptible *women* during the *1st & 2nd Time Periods*, $(Zw_1 \; and \; Zw_2)$, can be stated as:

$$F_w(a_1) = Zw_1 = P(MS, E \,\middle|\, F, G)_1 = \boldsymbol{d} * [1 - e^{-K(a_1)}] \qquad (Equation\ S5f)$$

and: $$F_w(a_2) = Zw_2 = P(MS, E \,\middle|\, F, G)_2 = \boldsymbol{d} * [1 - e^{-\{K(a_1) + \, q_w\}}] \qquad (Equation\ S5g)$$

where $\{K(a_1)\}$ indicates the *exposure-level* in *women* during the *1st Time Period* and the term $(q_w)$ is called the *"actual" exposure-level* change for *women* that has occurred between the two *Time Periods*. Therefore:

$$K(a_2) - K(a_1) = \; q_w > 0$$

Also, in a directly analogous manner to the derivation of *Equation S5e* (*above*):

$$q_w = \ln(1 - Zw_1/\boldsymbol{d}) - \ln\,(1 - Zw_2/\boldsymbol{d}) \qquad (Equation\ S5h)$$

Therefore, similar to those circumstances in susceptible *men*, the *"minimum"* possible value ($q_w^{min}$) for the *exposure-level* change in susceptible *women* will occur when: $(\boldsymbol{d} = 1)$, so that:

$$q_w^{min} = \ln(1 - Zw_1) - \ln\,(1 - Zw_2)$$

and: $$q_w \geq q_w^{min}$$

#### *5c. Relationship between Failure to True Survival*

In true survival everyone dies if given a sufficient amount of time. By contrast, as the *exposure-probability*, $P\big(E \,\big|\, G, E_T\big)$, approaches unity, the probability of failure (i.e., developing MS), either for susceptible *women* $(Zw)$ or for susceptible *men* $(Zm)$, may not approach 100%. Moreover, the *maximum* possible value for this *failure-probability* for susceptible *men* $(\boldsymbol{c})$ might not be the same as the *maximum* possible *failure-probability* for susceptible *women* $(\boldsymbol{d})$. Although the values of these $(\boldsymbol{c})$ and $(\boldsymbol{d})$ parameters are unknown, they are constants whenever the pathogenesis of disease involves environmental events, and regardless of whether the hazards are proportional. Finally, because exposure is being measured as the *odds* that the proband experiences a "*sufficient*" environment, the "*threshold*" exposure (i.e., the *exposure-level* at which MS becomes possible) must occur at: $P(E \,|\, G, E_T) = 0$; for susceptible *men*, or for susceptible *women*, or for both, provided that this *exposure-level* is possible [3]. If the hazards are proportional, the *threshold-difference* $(\lambda)$ is defined to be the difference between the threshold in susceptible *women* $(\lambda_w)$ and that in susceptible *men* $(\lambda_m)$ – i.e., $(\lambda = \lambda_w - \lambda_m)$. Consequently, if the threshold in *men* is greater than that in *women*, $(\lambda)$ will be negative and $(\lambda_w = 0)$; if the threshold in *women* is greater than that in *men*, $(\lambda)$ will be positive and $(\lambda_m = 0)$; If the threshold in *women* and *men* is the same, then: $(\lambda = \lambda_w = \lambda_m = 0)$.

Also, in true survival, both the clock and the risk of death begin at time-zero and continue into the future indefinitely. As a consequence, the cumulative probability of death increases monotonically with time. By contrast, for MS, it may be that the prevailing environmental conditions, during some *Time Period* $(E_T)$, are such that: $P(E \mid G, E_T) = 0$; even for a very extended *Time Period* (e.g., for centuries or millennia). Moreover, unlike the cumulative probability of death, for MS, the *exposure-level* may vary in any direction with time, depending upon the specific environmental conditions during $(E_T)$. Therefore, despite the cumulative probability of failure (i.e., of developing MS) increasing monotonically with increasing *exposure-level*, it may decrease, increase, or stay constant with time.

#### *5d. Relationship of the (F:M) Sex Ratio to Exposure*

Regardless of $(\lambda)$, and regardless of whether the hazards are proportional, the *failure-probability* during any *Time Period* for susceptible *women* $(Zw)$ can be stated as:

$$Zw = P(MS, E \mid G, F, E_T) = P(E \mid G, F, E_T) * P(MS \mid E, G, F)$$

or: $$Zw = P(E \mid G, F, E_T) * \boldsymbol{d}$$

and, similarly, the *failure-probability* for susceptible *men* $(Zm)$ can be stated as:

$$Zm = P(MS, E \mid G, M, E_T) = P(E \mid G, M, E_T) * \boldsymbol{c}$$

Dividing the *1st* of these two *Equations* by the *2nd*, during any *Time Period*, yields:

$$Zw/Zm = [P(E \mid G, F, E_T)/P(E \mid G, M, E_T)] * [\boldsymbol{d}/\boldsymbol{c}] \qquad \textit{(Equation S5i)}$$

Consequently, during any *Time Period*, any disparity observed between $(Zw)$ and $(Zm)$, must be due to a difference between *men* and *women* in the likelihood of their experiencing a "*sufficient*" environmental exposure, to a difference in the values of constants $(\boldsymbol{c})$ and $(\boldsymbol{d})$, or to a difference in both.

Therefore, by assuming that: $(\boldsymbol{c} = \boldsymbol{d} \leq 1)$, one is also assuming that any difference observed in disease expression between susceptible *women* and *men* is due entirely to a difference between susceptible *men* and *women* in the likelihood of their experiencing a "*sufficient*" exposure, despite the fact that, for every $(i)$, the exposure $\{E_i\}$ is both fixed and *population-wide* during any $(E_T)$. Thus, this exposure is "*available*" to everyone, so that, if the "*sufficient*" *exposure-level* differs between sexes, one possible explanation might be a systematic behavioral difference between susceptible *women* and *men* – i.e., to an increased exposure to, or avoidance of, susceptible environments by one or the other sex (perhaps consciously or unconsciously; or perhaps as a result of differing recreational activities, differing occupations, differing gender-roles, etc.). Nevertheless, the fact that *most men* behave differently from *women* does not indicate that *all men* do so, which makes a difference in *threshold* difficult to rationalize. Notably, also, if a finding of $(\lambda \neq 0)$ were to be explained by a systematic behavioral difference, then the finding of $(\lambda > 0)$ would suggest that the behavior of *men* leads to a greater exposure than the behavior of *women*. Any general conclusion in this regard, however, cannot be easily rationalized with the *current* observation that: $(Zw_2 > Zm_2)$. – *see Section 3a* (*above*)*; see also Supplemental Material; Reference #4*.

Another possible explanation for $(\lambda > 0)$, is that there may be distributions of so-called "*critical exposure intensity*" levels (i.e., "*thresholds*") that differ between susceptible *men* and *women* who are members of the same "*i-type*" group (*see Supplemental Material; Reference #4*). In such a case, perhaps, despite the fact that the same "*exposure-level*" is experienced equally by the two sexes, the "*intensity*" of this exposure might be disproportionately "*sufficient*" for susceptible *women* or susceptible *men.*

Membership in $(G)$ is presumed independent of $(E_T)$ so that the proportion of *women* among susceptible individuals $[p = P(F \mid G)]$ is also independent of $(E_T)$. Following the logic and notation leading to *Equation S4h* (*Section 4b; above*), therefore, regardless of whether the hazards are proportional, for any solution, the observed (*F:M*) *sex ratio* during any *Time Period* will be proportional to the observed $(Zw/Zm)$ ratio. Thus:

$$(F{:}M)\ sex\ ratio = \frac{P(MS,F \mid E_T)}{P(MS,M \mid E_T)} = \left(\frac{Zw}{Zm}\right) * \left(\frac{p}{1-p}\right) \qquad (Equation\ S5j)$$

#### *5e. Response Curves to Increasing Exposure*

From *Section 5a* (*above*) the response curves for both susceptible *men* and *women* are exponential. Importantly, any two points on *any* exponential curve define the entire curve. Thus, the values of $Zw$, $Zm$, $P(MS)$, and the (*F:M*) *sex ratio* during *any* two *Time Periods* define these response curves in susceptible *men* and *women – see Equations S5a & S5b* and *S5f & S5g* (*above*). Moreover, if the curves for both sexes can be plotted on the same *x-axis* (i.e., if both sexes are responding to the same environmental events), the hazards are always proportional (*see Section 7h; below*). Also, in this circumstance, the values of $(R = q_w/q_m)$ and $(\lambda)$ are determined from *Equations S7f-g* (*below*).

### *6. Non-proportional Hazard Models*

#### *6a. General Considerations*

If the hazard functions for *men* and *women* are not proportional, the "*actual*' *exposure-level* changes for *men* and *women* could each be at their *"minimums"* – i.e., $(q_m^{min})$ and $(q_w^{min})$. Such a circumstance, however, occurs when, and only when: $(\boldsymbol{c} = \boldsymbol{d} = 1)$ – *see Section 5b* (*above*).

Also, in this circumstance, although the "*plausible*" *parameter-value-ranges* for both observed and non-observed epidemiological parameters (*see Table 2; Main Text*) still limit possible solutions and, although $(\boldsymbol{c} \leq 1)$ and $(\boldsymbol{d} \leq 1)$ will be constants, nothing about them or about their relationship to each other can be inferred from the changes that take place in the (*F:M*) *sex ratio* and $P(MS)$ over time. Thus, any differences in the *values* that these *parameters* take during different *Time-Periods* could be attributed, both potentially and plausibly, to the differing environmental circumstances of different times and different places. In this circumstance, both the hazard proportionality factor $(R)$ and the parameter $(\lambda)$ – which relates the threshold in susceptible *women* to that in susceptible *men* – are meaningless.

Nevertheless, during *any Time Period*, the ratio of $(Zw/Zm)$ will still be proportional to the observed (*F:M*) *sex ratio* (*see Equation S5j*) and, if: $\boldsymbol{c} = \boldsymbol{d} \leq 1$, then any observed difference between $(Zw)$ and $(Zm)$, *must* be the result of a difference between susceptible *women* and susceptible *men* in the likelihood that they have experienced a "*sufficient*" environmental exposure during that *Time Period* (*see Equation S5i*).

***7. Proportional Hazard Models***

***7a. General Considerations***

If the hazards for *women* and *men* are proportional with the proportionality factor $(R)$, the situation is altered. First, because $(R > 0)$, the *penetrance-values* of $P(MS \mid F, G)$ and $P(MS \mid M, G)$, if they change over time, *must* have the same directionality. Indeed, the epidemiological observation that *MS-prevalence* has been increasing for both *women* and *men* over the past several decades, accords with this requirement [3,4,23]. Second, for the proportional hazard *Model*, including the possibility of a difference in threshold between the sexes, the *exposure-level* for susceptible *women* is represented as:

$$\forall\, H(a) \geq \lambda: \quad K(a) = R * \{H(a) - \lambda\} \geq 0 \qquad (\textit{Equation S7a})$$

In this circumstance, *Equations S5f & S5g*, which represent the *failure-probabilities* during the *1st& 2nd Time Periods* for susceptible *women*, can be re-stated as:

$$Zw_1 = \boldsymbol{d} * \left[1 - e^{-K(a_1)}\right] = \boldsymbol{d} * [1 - e^{-R*\{(H(a_1)-\lambda\}}] \qquad (\textit{Equation S7b})$$

and: $$Zw_2 = \boldsymbol{d} * \left[1 - e^{-K(a_2)}\right] = \boldsymbol{d} * [1 - e^{-R*\{H(a_1)+q_m-\lambda\}}] \qquad (\textit{Equation S7c})$$

*Equations S5a & S7b* can be rearranged for *any Time Period* to yield:

$$1 - Zw/\boldsymbol{d} = e^{-K(a)} = e^{-R*\{H(a)-\lambda\}} \qquad (\textit{Equation S7d})$$

and: $$1 - Zm/\boldsymbol{c} = e^{-H(a)} \qquad (\textit{Equation S7e})$$

Dividing *Equation S7d* by *S7e*, this result can be rearranged to yield:

$$\lambda = \{\ln[1 - Zw/\boldsymbol{d}] - \ln[1 - Zm/\boldsymbol{c}]\}/R + [(R-1)/R] * H(a) \qquad (\textit{Equation S7f})$$

Then *Equation S7f* can be applied to the *exposure-levels* $H(a_1)$ and $H(a_2)$ and one can subtract the 2nd of the resulting two *Equations* from the 1st. Then, applying *Equations S5e & S5h*, together with the defining *Equations* for $(q_m)$ and $(q_w)$ from *Section 5b* (*above*), this result can be rearranged to yield:

$$(R-1) * (q_m) = (q_w - q_m)$$

or: $$R = q_w/q_m \qquad (\textit{Equation S7g})$$

In addition, under circumstances where: $(R = 1)$, *Equation S7f* becomes:

$$\lambda = \ln[1 - Zw/\boldsymbol{d}] - \ln[1 - Zm/\boldsymbol{c}] \qquad (\textit{Equation S7h})$$

At any specific *exposure-level* $[H(a) \geq \lambda]$, the values of $(Zw)$ and $(Zm)$ are unknown. However, if a proportional hazard *Model* is appropriate for the disease being considered, the parameters $(\boldsymbol{c}, \boldsymbol{d}, R, \&\ \lambda)$ are constants (albeit unknown), so that, from *Equations S7d & S7e*, the probabilities of $(Zm)$ and $(Zw)$ are also fixed at any specific *exposure-level* $[H(a)]$.

***7b. Defining an "Apparent" Proportionality Factor***

An "*apparent*" hazard proportionality factor $(R^{app})$ can be defined such that: $R^{app} = (q_w^{min}/(q_m^{min})$, which represents the value $(R)$ when: $(\boldsymbol{c} = \boldsymbol{d} = 1)$ – see *Section 6a*; *above*. Potentially, this value incorporates two different processes. First, it may reflect the increased level of "*sufficient*" exposure experienced by one sex compared to the other. Indeed, from *Equation S5i*, this is the only possible interpretation for circumstances in which: $(\boldsymbol{c} = \boldsymbol{d} \leq 1)$. Second, however, if: $(\boldsymbol{c} < \boldsymbol{d} \leq 1)$ is admitted as a possibility, then a portion of $(R^{app})$ will be due to the difference of $(\boldsymbol{c})$ from unity.

*{NB: The possibility that: $(\boldsymbol{d} < \boldsymbol{c})$, is analogous to that of: $(\boldsymbol{c} < \boldsymbol{d})$, and, thus, is not specifically addressed.}*

Considering those circumstances in which: $(\boldsymbol{d} = 1)$ & $(R \geq 1)$, from *Sections 5b* (*above*) and *Section 8a* (*below*), the *"actual" exposure-level* change in *men* $(q_m)$ has a limited range such that:

$$\forall(R^{app} \geq R \geq 1): \quad q_m^{min} \leq q_m \leq q_w^{min}$$

$$\text{where:} \quad \boldsymbol{c} = (Zm_2) * \{e^{q_m} - [P(M,MS)_1/P(M,MS)_2]\}/(e^{q_m} - 1) \leq 1$$

From this, the *"actual"* hazard proportionality factor $(R^{app} \geq R \geq 1)$, at $(\boldsymbol{d} = 1)$, can be defined such that:

$$R^{app} \geq R = q_w^{min} \,/\, q_m$$

In this manner, if $(q_m > q_m^{min})$, some of the "*apparent*" value $(R^{app})$ will be accounted for by the fact that, in this case, $(\boldsymbol{c} < 1)$. Furthermore, if a reduction of $(\boldsymbol{c})$ from unity is possible in susceptible *men*, then, clearly, it is also possible for the value of $(\boldsymbol{d})$ in susceptible *women* to be less than unity. For example, when: $(\boldsymbol{c} < \boldsymbol{d} < 1)$, the *"actual" exposure-level* in *women* $(q_w)$ will be greater than its *minimum* value $(q_w^{min})$ such that:

$$R = q_w/q_m > q_w^{min}/q_m$$

As a result, in each of these cases, the *"actual"* $(R)$ value may differ from its *"apparent"* value $(R^{app})$.

#### *7c. Implications that the Values of* $(\boldsymbol{R})$*,* $(\boldsymbol{\lambda})$*,* $(\boldsymbol{c})$ *and* $(\boldsymbol{d})$ *have for Each Other*

***Assertions:***

1. $\forall(R \geq 1): \lambda > 0$
2. $\forall(\lambda \leq 0): \boldsymbol{c} < \boldsymbol{d} \leq 1$
3. $\forall(R \leq 1)$ & $\forall(R < R^{app}): \boldsymbol{c} < \boldsymbol{d} \leq 1$
4. $\forall(\boldsymbol{c} = \boldsymbol{d} \leq 1): both\ (R > 1)\ and\ (\lambda > 0)$

***Proof:*** The ratios $(C_F$ & $C_M)$ are defined in *Section 8a* (*below)* and, because both $P(MS)$ and the (*F:M*) *sex ratio* are *currently* increasing [3,4,23] – *see also Section 10a; Figure S1* (*below*) – therefore:

$$C_F = P(F,MS)_1/P(F,MS)_2 < P(M,MS)_1/P(M,MS)_2 = C_M$$

From *Equation S5j*, during *any Time Period*:

$$(F{:}M)\ sex\ ratio = (Zw/Zm) * \{p/(1-p)\}$$

and, as noted earlier, $[p = P(F \mid G)]$ is independent of the environmental conditions during $(E_T)$. Therefore, for all solutions, the $(Zw/Zm)$ ratio mirrors the (*F:M*) *sex ratio* – *see Section 5d* (*above*).

1. *For those Conditions in which:* $(R = 1)$:

From *Section 7a* (*above*) for circumstances where: $\{R = (q_w/q_m) = 1\}$, it must be that:

$$q_m = q_w \geq q_w^{min}$$

When: $(\lambda = 0)$, from *Equation S7h* (*above*):

$$Zm/\boldsymbol{c} = Zw/\boldsymbol{d}$$

or: $$Zw/Zm = \boldsymbol{d}/\boldsymbol{c}$$ (*Equation S7i*)

Therefore, the *F:M sex ratio* will remain constant in this case, regardless of the *exposure-level*.

However, because in this case: $[q_w = q_m]$; therefore, from the *Equations S8c-d* (*below*):

$$\boldsymbol{d}/\boldsymbol{c} = \{Zw/Zm\} * \{(e^{q_w} - C_F)/(e^{q_w} - C_M)\} > Zw/Zm$$

or, with rearrangement: $Zm/\boldsymbol{c} > Zw/\boldsymbol{d}$

Therefore, from *Equation S7h*: $\lambda > 0$

Consequently, if $(R = 1)$, and if both the *F:M sex ratio* and $P(MS)$ are *currently* increasing, then the threshold for susceptible *women* <u>must</u> be greater than that it is for susceptible *men*.

2. *For those Conditions in which:* $(\lambda \leq 0)$ & $(R > 1)$:

For $\{H(a) \geq 0\}$, from *Equation S7f*, during any $(E_T)$, under these conditions:

$$\{\ln(1 - Zw/\boldsymbol{d}) - \ln(1 - Zm/\boldsymbol{c})\}/R = \lambda - [(R-1)/R] * H(a) \leq 0$$

or: $$\ln(1 - Zw/\boldsymbol{d}) - \ln(1 - Zm/\boldsymbol{c}) \leq 0$$ (*Equation S7j*)

In turn, under these conditions, *Equation S7j* requires that:

$$Zm/\boldsymbol{c} \leq Zw/\boldsymbol{d}$$

or: $$Zw/Zm \geq \boldsymbol{d}/\boldsymbol{c}$$ (*Equation S7k*)

Also, regardless of the value of $(R)$, from the definitions of $(\boldsymbol{c})$, and $(\boldsymbol{d})$ – *Section 5b* – from the definition of $(E)$ – *Section 5b* – and from *Equation S5i*:

$$\lim_{a \to \infty}(Zw/Zm) = \boldsymbol{d}/\boldsymbol{c}$$ (*Equation S7l*)

Because, with increasing exposure, both $(Zw)$ and $(Zm)$ increase monotonically (*see Section 4a*), and because $(R > 1)$, and because $(\lambda \leq 0)$, and because $\{H(a) \geq 0\}$, the condition that:

$$Zw/Zm \geq \boldsymbol{d}/\boldsymbol{c}$$

requires that: $Zw_1/Zm_1 \geq Zw_2/Zm_2 \geq \boldsymbol{d}/\boldsymbol{c}$:

Thus, under these conditions, the $(Zw/Zm)$ ratio either decreases or remains constant with increasing exposure. Because the $(Zw/Zm)$ ratio mirrors the *F:M sex ratio*, therefore, the *F:M sex ratio* will also decrease or remain constant (e.g., *Figure 1C; Main Text*) – a conclusion, which is inconsistent with the evidence [1-4,23]. Thus, the conditions: $(R > 1)$ & $(\lambda \leq 0)$ are not plausible, given the Canadian data [23].

Combining these two conclusions (i.e., *Conditions 1 & 2* ; *above*), it *must* be the case that:

$$\forall(R \geq 1)\colon\ \lambda > 0$$

From the Canadian MS data [23], both $P(MS)$ and the (*F:M*) *sex ratio* are *currently* increasing when the "*current*" epoch is compared to any of the previous 5-year epochs from the same study – *see Section 10a, Figure S1* (*below*). An increasing MS-prevalence disproportionately affecting *women* is also reported from other parts of the world [1-4]. Therefore, based exclusively on the increasing $P(MS)$ and (*F:M*) *sex ratio*, and on purely theoretical grounds, one can conclude, that, if the hazards in *men* and *women* are proportional and if: $(R \geq 1)$, then susceptible *women* *must* have a higher threshold than susceptible *men*.

3. *For those Conditions, in which:* $(\lambda \geq 0)$ & $(R \leq 1)$

If: $(\lambda \geq 0)$ & $(R \leq 1)$ & $(\boldsymbol{c} = \boldsymbol{d} \leq 1)$; then the *failure-probability* for susceptible *men* would be as great (or a greater) than the *failure-probability* for *women* (i.e., $Zm \geq Zw$) at every *exposure-level* (*see Figure 2B; Reference #4*). Because: $(Zw_2 > Zm_2)$, these conditions are impossible. Therefore, whenever: $(\lambda \geq 0)$ & $(R \leq 1)$, then: $(\boldsymbol{c} < \boldsymbol{d} \leq 1)$ – e.g., *Figure 1B* (*Main Text*).

4. *For those Conditions, in which:* $(\lambda < 0)$ & $(R \leq 1)$:

In these conditions, *Equation S7k* still applies and, thus, if: $(\boldsymbol{c} = \boldsymbol{d} \leq 1)$, following the intersection of the response curves for susceptible *men* and *women*, then $(Zm > Zw)$ at every *exposure-level* (e.g., *Figure 1; Main Text*). Because an increasing *F:M sex ratio* only takes place after this intersection, the condition that both: $(Zw_2 > Zm_2)$ & $(\boldsymbol{c} = \boldsymbol{d} \leq 1)$, is not possible. Nevertheless, the condition that: $(\boldsymbol{c} < \boldsymbol{d} \leq 1)$ is still possible – e.g., *Figure 1D* (*Main Text*). Therefore, combining *Conditions 2 & 4* (*above*), it *must* be the case that:

$$\forall(\lambda \leq 0)\colon\ \boldsymbol{c} < \boldsymbol{d} \leq 1$$

5. *For those Conditions, in which:* $(R^{app} > 1)$ *or* $(R^{app} > R)$:

The value of $(R)$ is related to how quickly the response curves for *men* and *women* go from onset to their maximums. Thus, this value is independent of $(\lambda)$. Rather, it depends only upon how quickly this transition occurs. Consequently, for comparison, one is free to choose *any* $(\lambda)$ value. Therefore, when $(\boldsymbol{c} = \boldsymbol{d})$ & $(\lambda = 0)$, for any $(E_T)$, *Equations S5a & S5f* can be multiplied by the scaling factor of: $(1/\boldsymbol{c})$, and then restated as:

$$Zm/\boldsymbol{c} = (1 - e^{\{H(a)\}})$$

and: $$Zw/\boldsymbol{c} = (1 - e^{R*\{H(a)\}})$$

The *RHS* of both *Equations* is independent of scale. Also, the relationship *between* the *LHS* of two *Equations* is also independent of scale. Therefore, the relationship *between* these two *Equation*, when $(\boldsymbol{c} = \boldsymbol{d})$, is independent of scale. In his case, when: $(\boldsymbol{c} = \boldsymbol{d})$ the value of $(R)$ is constant for all: $(Zm_2 < \boldsymbol{c} \leq 1)$ and therefore:

$$\forall(\boldsymbol{c} = \boldsymbol{d})\colon\ R^{app} = q_w^{min}/q_m^{min} = q_w/q_m = R$$

However, whenever: $(R \leq 1)$, then also, $(q_w \leq q_m)$.

Consequently, whenever: $(R^{app} > 1)$, then:

$$R^{app} = \ q_w^{min}/q_m^{min} > 1 \geq q_w/q_m = R$$

Any circumstance in which: $(R^{app} > 1)$, therefore, implies that:

$$\forall(R \leq 1): \ \ \boldsymbol{c} < \boldsymbol{d}$$

Combining the three conclusions from *Conditions 3–5* (*above*), it is clear that:

$$\forall(R \leq 1): \ \ \boldsymbol{c} < \boldsymbol{d} \leq 1$$

Indeed, following a logic directly analogous to that *above*, it must also be that:

$$\forall(R^{app} > R): \ \boldsymbol{c} < \boldsymbol{d}$$

6. <u>*Finally:*</u> Combining each of the conclusions from *Conditions 1–5* (*above*), one can further conclude, based on purely theoretical grounds, that whenever: $(\boldsymbol{c} = \boldsymbol{d} \leq 1)$, it <u>*must*</u> also be the case that both: $(R > 1)$ and: $(\lambda > 0)$.

***7d. Strictly Proportional Hazard:*** $(\boldsymbol{\lambda} = \mathbf{0})$

If the condition of "strictly" proportional hazards in *men* and *women* were to apply, then, by definition: $(\lambda = 0)$. Consequently, whenever: $(\lambda > 0)$, as it <u>*must*</u> be when $(R \geq 1)$, the hazards cannot be "strictly" proportional to each other. In fact, for those cases in which $(R \geq 1)$ and $(\lambda = 0)$, the observed (*F:M*) *sex ratio* either decreases or remains constant with increasing exposure (*see Equations S7j–l; above*), regardless of the values that $(\boldsymbol{c})$ and $(\boldsymbol{d})$ parameters take – e.g., *Figure 1C* (*Main Text*). Therefore, the only possible "strictly" proportional conditions, are those in which the hazard in susceptible *men* is greater than that in susceptible *women* – i.e., $(R < 1)$. Importantly, if the hazard in *men* is greater than that in *women*, then, as noted in *Section 7c;* (*above*) the simultaneous conditions of: $(\boldsymbol{c} = \boldsymbol{d} \leq 1)$ & $(\lambda = 0)$ are excluded.

Consequently, the only "strictly" proportional conditions possible are those, in which both $(R < 1)$ and $(\boldsymbol{c} < \boldsymbol{d} \leq 1)$ – e.g., *Figure 1D* (*Main Text*).

*{NB: In the Figures presented in the Main Text, all response curves serving as examples for conditions in which:* $(\boldsymbol{c} = \boldsymbol{d} \leq 1)$, *are depicted for the condition* $(\boldsymbol{c} = \boldsymbol{d} = 1)$. *Nevertheless, for all conditions (and, therefore, for all Figures) in which the condition of* $(\boldsymbol{c} = \boldsymbol{d} \leq 1)$ *applies, the depicted response curves differ only in so far as the scale of the y-axis is different. Thus, any response curve, depicted at:* $(\boldsymbol{c} = \boldsymbol{d} = 1)$, *is representative of all curves for conditions in which* $(\boldsymbol{c} = \boldsymbol{d})$ – *see Section 7c; Condition 5 (above).}*

***7e. Intermediate Proportional Hazard:*** $(\boldsymbol{\lambda} < \mathbf{0})$

It is possible that a different *Model*, the so-called "*intermediate*" *Model*, is more appropriate than the "strictly" proportional *Model* considered *above*. In this *Model*, the hazards in susceptible *women* and *men* are still held to be proportional to one another but the onset of the response curves in *women* and *men* are offset from each other by an amount $(\lambda \neq 0)$. As noted previously: $\forall\ (R \geq 1)$: $\lambda > 0$. Consequently, whenever: $(\lambda < 0)$, it *must* be that the hazard in *men* is greater than it is in *women*. In addition, under conditions, where $(\boldsymbol{c} = \boldsymbol{d} \leq 1)$ & $(R < 1)$ & $(\lambda < 0)$, the (*F:M*) *sex ratio* initially decreases with increasing exposure until the two response curves intersect at a point below $[p/(1-p)]$ on the *y-axis* (e.g., *Figure 1A; Main Text*). Following this intersection, the (*F:M*) *sex ratio* increases steadily, ultimately reaching a level of $[p/(1-p)]$ on the *y-axis* and, notably, never exceeds this level. In addition, after this intersection (i.e., after the nadir), the response curves maintain a relationship such that: $(Zm > Zw)$, throughout the remainder of response curve until the (*F:M*) *sex ratio* reaches the level of: $[p/(1-p)]$ *on the y-axis* (e.g., *Figure 1A; Main Text*). Moreover, defining the term: $[(p') = P(F \mid MS)]$; it follows from *Equation S5j* (*above*) and the condition that: $(Zw_2 > Zm_2)$ requires both of the conditions:

$$(p')_2 > p \qquad \& \qquad [p'/(1-p')]_2 > p/(1-p)$$

Therefore, the condition of: $(\lambda < 0)$ is only possible, when: $(\boldsymbol{c} < \boldsymbol{d})$ – e.g., *Figure 1B* (*Main Text*).

#### *7f. Intermediate Proportional Hazard:* $(\boldsymbol{\lambda} > \mathbf{0})$ *& Autosomal Genotypes*

By contrast, when $(\lambda > 0)$, there are no constraints on the relationship that the hazards can take in susceptible *women* compared to susceptible *men*. Thus, both the conditions of: $(R < 1)$ & $(\lambda \geq 0)$ and the conditions of: $(R \geq 1)$ & $(\lambda > 0)$ lead to similar conclusions (*see Figures 3 & 4; Reference #4*).

In this case, it is useful to define a so-called "*susceptibility genotype*", $(G_{is})$, for the $i^{th}$ susceptible individual. This genotype includes only those genetic factors (located on any chromosome), which are related to MS susceptibility. Because $(G_{is})$ includes the specification of fewer genetic factors than does the complete genotype of the $i^{th}$ individual $(G_i)$, it is possible for more than one person in the population to belong to the same *susceptibility-genotype*. For example, because *MZ*-twins have "*identical genotypes*", therefore, based on our assumption (*see Section 1a, above*), they necessarily have the same *susceptibility-genotype*. The group of individuals, who have the same *susceptibility-genotype* as the $i^{th}$ individual is referred to as the $(G_{is})$ subset within $(Z)$. The occurrence of $(G_{is})$ represents the event that a person, randomly selected from $(Z)$, belongs to the $(G_{is})$ subset. The probability of this event is represented as $P(G_{is})$. Because some members of $(G)$ are *MZ*-twins, therefore, the total number of these *susceptibility-genotypes* in the population $(m_{is})$ is less than $(m)$ – i.e., $(m_{is} < m)$. The subset $(G_s)$ includes *all* of the *susceptibility genotypes* within $(Z)$. The occurrence of $(G_s)$ represents the event that an individual, selected randomly from $(Z)$, is member of the $(G_s)$ subset.

Also, it is possible for two or more individuals (perhaps, each with a different *susceptibility genotype*) to share the same family of "*sufficient*" environmental exposures $\{E_i\}$ with the $i^{th}$ individual (*see Section 1a*). Therefore, the "*i-type*" *exposure-group* $(G_{it})$ – or the "*i-type*" group – is defined to include *all* individuals (possibly with different "*susceptibility genotypes*") who share the same $\{E_i\}$ family. The probability: $P(G_{it})$

represents the probability of the event, $(G_{it})$, that an individual, randomly selected from $(Z)$, belongs to the $(G_{it})$ *exposure-group*. Also, from *above*, the total number of "*i-type*" *exposure-groups* in the population $(m_{it})$ must be less than $(m)$ – i.e., $(m_{it} \leq m_{is} < m)$. The family $\{G_t\}$ is defined to include <u>*all*</u> of the "*i-type*" *exposure-groups*, $(G_{it})$, within $(Z)$.

The "*autosomal susceptibility genotype*" of the *i^th^* susceptible individual, $(G_{ia})$, is defined to include all of genetic factors (located on autosomal chromosomes) that are related to MS susceptibility. The occurrence of $(G_{ia})$ represents the event that an individual, randomly selected from $(Z)$, is a member is a member of the $(G_{ia})$ subset – a subset consisting of a single *autosomal susceptibility genotype*. The subset $(G_a)$ is defined to include all of these *autosomal susceptibility genotypes* within the $(G)$ subset. In a similar manner, the occurrence of $(G_a)$ represents the event that an individual, randomly selected from $(Z)$, is a member of the $(G_a)$ subset.

Because the genotypes within $(G_a)$ are exclusively autosomal, it is anticipated that:

$$\forall\, G_{ia} \in (G_a): \quad P(G_{ia} \mid M) = P(G_{ia} \mid F)$$

$$\forall\, G_{ia} \in (G_a): \quad P(G_{ia}, F, G, G_{is}) = P(F, G_{is})$$

and: $$\forall\, G_{ia} \in (G_a): \quad P(G_{ia}, M, G, G_{is}) = P(M, G_{is})$$

Certainly, it is possible for susceptible *women* and *men* may be members of the same $(G_{ia})$ subset, but not be members of the same $(G_{is})$ subset. Consequently, these anticipated equivalences do not, necessarily, imply either that:

$$\forall\, G_{is} \in (G_s): \quad P(F, G_{is}) = P(M, G_{is})$$

or that both: $\forall\, G_{is} \in (G_s): \quad P(F, G_{is}) > 0$ and: $\forall\, G_{is} \in (G_s): \quad P(M, G_{is}) > 0$

However, all but one of the 233 MS-associated genetic loci, reported by the *International Multiple Sclerosis Genetics Consortium*, are located on autosomal chromosomes [6]. Moreover, even for the single locus found on the X-chromosome, *men* and *women* both carried the risk-variant [6]. In such a circumstance, therefore, it seems very likely that:

$$\forall\, G_{is} \in (G_s): \quad P(F, G_{is}) \approx P(M, G_{is})$$

And that the same conclusion will hold for all "*i-type*" *exposure-groups* $(G_{it})$. Therefore, likely:

$$\forall\, G_{it} \in \{G_t\}: \quad P(F, G_{it}) \approx P(M, G_{it})$$

As a result, likely, both *men* and *women* (at least potentially) could belong to any of the "*i-type*" *exposure-groups* – in which case they will be referred to as "*i-type*" individuals. The same conclusion is suggested by the evidence from the occurrence of MS wthin families (*see Main Text*). In this context, those environmental factors, which comprise each of the "*sufficient*" *exposure-sets* within the $\{E_i\}$ family, are envisioned to be the same regardless of whether the "*i-type*" individual is a *woman* or a *man*. However, it may be that the "*sufficient*" exposure for an "*i-type*" *woman* needs to be more or less "*intense*" than it is for an "*i-type*" *man* [4].

*7g Considerations of Exposure "Intensity"*

Before considering notions of "*exposure-intensity*", it is notable that there seem to be four well-established conclusions. First, for *every* proportional hazard solution, which was identified by this analysis (*see Results; Main Text*), it was found that:

$$(R^{app} > 1)$$

Second, on theoretical grounds, from *Section 7c* (*above*), it <u>*must*</u> be the case that:

$$\forall(R \leq 1)\ \&\ \forall(R < R^{app})\ \ \&\ \forall(\lambda \leq 0):\ \ \boldsymbol{c} < \boldsymbol{d}$$

Third, from *Section 7c* (*above*), under those conditions where both $P(MS)$ and $P(F \mid MS)$ are increasing, then it <u>*must*</u> be the case that:

$$\forall(R \geq 1):\ \lambda > 0$$

And fourth, from the Canadian MS-data [23], as the probability of a "*sufficient*" environmental exposure has increased over the last several decades, so too has the (*F:M*) *sex ratio – see Sections 8a & 10a-b; see also Figure S1* (*below*). From these two observations, one can conclude that, over this period of time, the probability $[Zw = (MS \mid F, G)]$ <u>*must*</u> have increased at a faster rate than has the probability $[Zm = (MS \mid M, G)]$ and, therefore, almost certainly, it is *currently* the case that: $(Zw > Zm)$ *– see Section 3a* (*above*).

From these four conclusions, if susceptible *men* and *women* have proportional hazards, it follows (*see Section 7c; above*) that following three conditions <u>*must*</u> also hold.

1) if: $R \leq 1$ or if: $R < R^{app}$ or if: $\lambda \leq 0$ ; then: $\boldsymbol{c} < \boldsymbol{d}$
2) if: $\boldsymbol{c} = \boldsymbol{d} \leq 1$ ; then, both: $R > 1$ and: $\lambda > 0$
3) if: $R > 1$ ; then: $\lambda > 0$

Condition *#1*, clearly, excludes *any* possibility that: $\boldsymbol{c} = \boldsymbol{d} = 1$

Considering conditions *#2&3*, notably, both of the exposure measures used in this analysis – i.e., $(a)$ and $H(a)$ – are directly related to the parameter $P(E \mid G)$, which represents the probability of the event that an individual, randomly selected from the $(G)$ subset, experiences an environmental exposure "*sufficient*" to cause MS in them. Consequently, these conditions – i.e., where: $\lambda > 0$ – indicate that, as the odds of a "*sufficient*" exposure decreases, there must come a point where only susceptible *men* can develop MS. This implies that, at (or below) this *exposure-level*, $(R = 0)$. As a result, the additional requirement that: $(R > 1)$ poses a potential paradox in that, if either of these conditions were true, susceptible *women* would be more likely than *men* to experience a "*sufficient*" exposure when the probability $[P(E \mid G)]$ is high and, yet, susceptible *men* would be considerably more likely than *women* to experience a "*sufficient*" exposure when this probability is low.

There are two obvious ways to avoid this paradox. Principal among them is for one to conclude that the hazards are not proportional. Despite this possibility, however, such a conclusion creates other problems (*see Main Text*). For example, susceptible *women* and *men* who are members of the same "*i-type*" *exposure-group* necessarily have proportional hazards (*see Section 7h; below*). Therefore, in this case, one would also have to conclude further that susceptible *women* and *men* can never be in the same "*i-type*" *exposure-group* and, consequently, that the "*sufficient" exposure-sets* are different for the two sexes. In such a circumstance, MS in *women* would represent a different disease from MS in *men*. Alternatively, if it were possible that both *women* and *men* could be members of some "*i-type*" *exposure-groups* but not others, one would conclude that MS represents three distinct diseases (one in *women*, one in *men*, and a third in both). Neither conclusion is supported by the available genetic and the epidemiological evidence (*see Main Text*).

The second way to avoid the paradox is to accept *Condition #1*, which is compatible with any $(\lambda)$. However, if: $(\lambda > 0)$ and $(R \leq 1)$, then, at every population *exposure-level* $(a)$, the probability of the event that a *susceptible-man*, randomly-selected, will experience a *sufficient-exposure* is as great, or greater, than the same probability for a *susceptible-woman*. Thus, although developing a notion of a so-called "*critical exposure-intensity*" may be necessary to rationalize any threshold difference between susceptible *women* and *men* [4], it is not necessary to resolve any paradox. Nevertheless, accepting the conclusion that $(\lambda > 0)$ and $(R \leq 1)$, does require also accepting the fact that $(\boldsymbol{c} < \boldsymbol{d})$ and therefore that some susceptible *men* will never develop MS, even when the correct genetic background occurs together with an environmental exposure "*sufficient*" to cause MS in them (*see Section 7c; above*).

#### *7h. Variability in the Values of $(R_i)$ and $(\lambda_i)$ between "i-type" Groups*

In the circumstance where both *men* and *women* are (or potentially could be) members of some specific "*i-type*" *exposure-group* $\{G_{it}\}$, by definition, such *men* and *women* each will have *some* non-zero probability of developing MS in response to *every* "*sufficient*" *exposure-set* within the $\{E_i\}$ family. For notational clarity, a subset $(G_w)$ will be defined to include of <u>*all*</u> *female* members of the $(G)$-subset $\{$i.e., $(G_w) = (F, G)\}$. As in previous *Sections*, the proportion of *women* in the $(G)$ subset is defined as: $[p = P(F \mid G)]$. In this case, each of the $(m * p)$ *women* in the $(G_w)$ subset $(d = 1, 2, \ldots, mp)$ has a unique genotype $(G_w)$. The occurrence of $(G_{dw})$ represents the event that an individual, selected randomly from the population $(Z)$, belongs to the $(G_w)$ subset – a subset consisting of only single individual (i.e., the $d^{th}$ susceptible *woman*) – and the probability of this event is represented as: $\{P(G_{dw}) = 1/N\}$. Also, the probability of the event that an individual, selected randomly from the population $(Z)$, belongs to the $(G_w)$ subset is represented as: $\{P(G_w) = P(F, G) = mp/N\}$.

*{NB: The use of $(G_w)$ and $(G_{dw})$ terminology is used only when the listing of individual susceptible genotypes for women is important to the argument being made.}*

In the circumstance where both *men* and *women* are (or, potentially, could be) members of *every* "*i-type*" *exposure-group*, then, at every *exposure-level* for a *man* $\{H(a) \geq \lambda\}$, a proportionality constant $(R_i > 0)$ is defined, so that the *exposure-level* for any *i-type* susceptible *woman* $\{K_i(a) \geq 0\}$ can be stated as:

$$\forall\ G_{dw} \in (F, G_{it}):\ \ K_i(a) = R_i * \{H(a) - \lambda\}$$

*{NB: In this case, one doesn't need to consider the "i-type" specific exposure for men,* $H_i(a)$*, because, by definition, if each exposure-group has the same threshold difference* $(\lambda > 0)$ *then, for all* $\{H(a) \geq \lambda\}$ *and for all* $(i)$*, it will be true that, for all* $(a)$*, both* $\{H(a) - \lambda \geq 0]\}$ *and* $\{K_i(a) \geq 0\}$*. Consequently, in this case, there will be some constant* $(R_i > 0)$ *that permits this statement to be true for each* $(i)$*. The impact of different i-type exposure-groups having different thresholds is considered below.}*

Because each susceptible *woman* $(G_{dw})$ is a member of *some* "*i-type*" *exposure-group* $(G_{it})$, an *exposure-level* $[K_{dw}(a)]$ and a proportionality factor $[R_{dw}]$ can be defined for each susceptible *woman* so that:

$$\forall\ G_{dw} \in (G_w):\ \ K_{dw}(a) = R_{dw} * (H(a) - \lambda)$$

where: $\forall\ G_{dw} \in (F, G_{it}):\ K_{dw}(a) = K_i(a)\ \ and:\ \ R_{dw} = R_i$

In this circumstance, the expected *exposure-level* for susceptible *women* can be stated as:

$$K(a) = E\{K_{dw}(a)\} = \sum_{d=1}^{mp} R_{dw} * \{H(a) - \lambda\}/mp = R * \{H(a) - \lambda\}$$

where: $R = E(R_{dw})$

Consequently, if *women* and *men* can (at least potentially) be members of every "*i-type*" *exposure-group*, the hazards for *women* and *men* will always be proportional. However, the hazard proportionality factor $(R_i)$ may be different for different "*i-type*" *exposure-groups*.

It is also possible that the threshold-difference between *women* and *men* $(\lambda_i)$ varies between the different "*i-type*" *exposure-groups*. Initially, the circumstances where $(\lambda > 0)$ are considered. The "*i-type*" *exposure group* $(j)$ with the smallest threshold $(\lambda_{jm})$, for *men* of any "*i-type*" *group*, can be defined such that:

$$[\lambda_{jm} = \min(\lambda_m)]$$

By definition: $(\lambda_{jm} = 0)$ – *see Section 5c; above*. Similarly, in this case, the *i-type*" *exposure-group* $(k)$ with the smallest threshold $(\lambda_{kw})$, for *women* of any "*i-type*" *group* can be defined such that:

$$[\lambda_{kw} = \min(\lambda_w) > 0]$$

In this case, from the definition of threshold, some *men* and some *women* will begin to develop MS at these *exposure-levels* so that, in this circumstance:

$$\lambda = \lambda_{kw} - \lambda_{jm} = \lambda_{kw}$$

Moreover, it is possible that the *men* and *wome*n who develop MS at these *exposure-levels* are not members of the same "*i-type exposure-group* and, therefore, it is not necessarily the case that $(j = k)$. Regardless, however, a difference in threshold can then be defined between $(\lambda_{dw})$ for each susceptible *woman* and $(\lambda_{jm} = 0)$. In this circumstance, therefore, one can define one can define $(\lambda_i > 0)$ such that:

$$\forall\, G_{it} \in \{G_t\} \;\&\; \forall\, G_{dw} \in (F, G_{it}):\;\; \lambda_{dw} = \lambda_i$$

In this way, the proportionality constants for each "*i-type*" $(R_i > 0)$ and each *woman* $(R_{dw} > 0)$ can be replaced by a *"adjusted"* proportionality constants $(R'_i > 0)$ and $(R'_{dw} > 0)$ such that:

$$\forall\, G_{dw} \in (G_w):\;\; K_{dw}(a) = R_{dw} * (H(a) - \lambda_{dw}) = R'_{dw} * \{H(a) - \lambda\}$$

where: $\forall\, G_{dw} \in (F, G_{it})$: $K_{dw}(a) = K_i(a)$ ; $R_{dw} = R_i$ ; $R'_{dw} = R'_i$ ; and: $\lambda_{dw} = \lambda_i$

Thus, in this circumstance, the expected *exposure-level* for susceptible *woman* can be stated as:

$$K(a) = E\{K_{dw}(a)\} = \sum_{d=1}^{mp} R'_{dw} * \{H(a) - \lambda\}/mp = R * \{H(a) - \lambda\}$$

where: $R'_{dw} = R_{dw} * \{(H(a) - \lambda_{dw})/(H(a) - \lambda)\} \leq R_{dw}$

and where now: $R = E(R'_{dw})$

When: $(\lambda < 0)$, this analysis is only changed in that the roles of susceptible *men* and *women* are interchanged for all of the above arguments and conditions. Thus, in both cases, the hazards will be proportional. Moreover, because *failure-probability* is described only as a function of the *probability* of a "*sufficient*" exposure, given the environmental conditions of the time (*see Section 5a; above*), and because it is posited that *women* and *men* can (at least potentially) be members of every "*i-type*" *exposure-group*, it is unnecessary to specify the composition of the "*sufficient*" *exposure-sets*, within each $\{E_i\}$, which have resulted in the observed *failure-probability* change between *Time Period #1* and *Time Period #2*.

By contrast, if *men* and *women* each require distinct "*sufficient*" *exposure-sets*, the hazards will not be proportional and *women* and *men* would require their response curves plotted separately; each graph having its own *x-axis* scale. In this case, one would also need to envision *men* and *women* with MS as each having different underlying diseases.

*{NB: One might also imagine the possibility that $(R_i)$ or $(\lambda_i)$ or both varied between the different exposure-sets within $\{E_i\}$. In such a circumstance, susceptible-men and susceptible-women (considered separately) would still have an exponential relationship between their failure-probability and their exposure as measured by the odds that a proband (either male or female) experiences an exposure "sufficient" to cause MS in them (see Section 5a; above). However, if this variability were large enough, the relationship between "i-type" men and "i-type" women could become non-proportional and effectively equivalent to those circumstances, in which these men and women were actually members of distinct "i-type" exposure-groups. In this case, for such "i-type" individuals, as is also the case in other non-proportional circumstances (see above), female-MS and male-MS would represent distinct diseases.}*

#### 8. *Summary Equations for the Longitudinal Model*

##### *8a. Derivations*

For notational simplicity, three related ratios are defined:

$$C = P(MS)_1/P(MS)_2 \quad \text{or:} \quad P(MS)_1 = C * P(MS)_2$$

$$C_F = P(F,MS)_1/P(F,MS)_2 = C * [P(F \mid MS)_1/P(F \mid MS)_2]$$

$$C_M = P(M,MS)_1/P(M,MS)_2 = C * [P(M \mid MS)_1/P(M \mid MS)_2]$$

The following *Summary Equations* can be derived using these definitions:

1. First, one can re-express $(Zw_2)$ & $(Zw_1)$ such that:

$$Zw_2 = P(MS,E \mid G,F)_2 = P(MS \mid G,F)_2 = P(F \mid MS)_2 * \left(\frac{P(MS)_2}{P(G,F)}\right)$$

$$Zw_1 = P(MS \mid G,F)_1 = \frac{P(MS)_1 * P(F \mid MS)_1}{P(G,F)} = C * P(F \mid MS)_1 * \left(\frac{P(MS)_2}{P(G,F)}\right)$$

Therefore: $Zw_2/P(F \mid MS)_2 = Zw_1/\{C * P(F \mid MS)_1\}$

so that: $$Zw_1 = Zw_2 * C * \left(\frac{P(F \mid MS)_1}{P(F \mid MS)_2}\right) = Zw_2 * \left(\frac{P(F,MS)_1}{P(F,MS)_2}\right) = Zw_2 * C_F \qquad \textit{Equation S8a}$$

and similarly: $$Zm_1 = Zm_2 * C * \left(\frac{P(M \mid MS)_1}{P(M \mid MS)_2}\right) = Zm_2 * \left(\frac{P(M,MS)_1}{P(M,MS)_2}\right) = Zm_2 * C_M \qquad \textit{Equation S8b}$$

*Equation S5d* (*see Section 5b; above*) for *men* can then be rearranged to yield:

$$\boldsymbol{c} = \{e^{q_m} * Zm_2 - Zm_1\}/(e^{q_m} - 1)$$

Substituting in this equation for $(Zm_1)$ from *Equation S8b* yields:

$$\boldsymbol{c} = Zm_2(e^{q_m} - C_M)/(e^{q_m} - 1) \qquad \textit{Equation S8c}$$

and similarly: $$\boldsymbol{d} = Zw_2(e^{q_w} - C_F)/(e^{q_w} - 1) \qquad \textit{Equation S8d}$$

2. Also, notably, both: $(Zm_2 < \boldsymbol{c})$; and: $(Zw_2 < \boldsymbol{d})$. Therefore, from *Equation S8c* and from the definition of the ratio $(C_M)$ – *see above* – it must be the case that:

$$Zm_2 < Zm_2 * \{e^{q_m} - C * \{P(M \mid MS)_1/P(M \mid MS)_2\}/(e^{q_m} - 1)$$

Dividing both sides of this inequality by $(Zm_2)$ and, with rearrangement, yields:

$$C < P(M \mid MS)_2/P(M \mid MS)_1 \qquad \textit{Equation S8e}$$

and similarly: $$C < P(F \mid MS)_2/P(F \mid MS)_1 \qquad \textit{Equation S8f}$$

One can use the point estimates from *Section 10b* (*below*) – i.e., $P(M \mid MS)_2 = 0.238$ & $P(M \mid MS)_1 = 0.315$ – and, inserting these estimates into *Equation S8e*, yields:

$$C < P(M \mid MS)_2 / P(M \mid MS)_1 = 0.238/0.315 = 0.756$$

Therefore, the observations from the *CCPGSMS* dataset [23] translate to a *minimum* increase in *MS-penetrance* by more than 32% between *Time Period #1* and *Time Period #2* – or, equivalently, to an increase in the prevalence of MS in Canada by more than 32% between the two *Time Periods*.

3. And, finally, because: $P(MS \mid E, G, M) = \boldsymbol{c}$ and: $P(MS \mid E, G, F) = \boldsymbol{d}$ ; during any *Time Period*, then:

$$Zm_2 = P(MS, E \mid G, M)_2 = P(E \mid G, M)_2 * P(MS \mid E, G, M)$$

or: $$Zm_2 = P(E \mid G, M)_2 * (\boldsymbol{c})$$

with rearrangement, this becomes:

$$P(E \mid G, M)_2 = P(MS, E \mid G, M)_2 / \boldsymbol{c}$$ *Equation S8g*

and, similarly: $$P(E \mid G, F)_2 = P(MS, E \mid G, F)_2 / \boldsymbol{d}$$ *Equation S8h*

#### *8b. Limits on the Value of the Parameters:* $\boldsymbol{P(MS \mid E)}$, $(\boldsymbol{c})$ *and* $(\boldsymbol{d})$

As noted earlier (*see Section 2a*), the observed *MZ*-twin concordance rate [i.e., $P(MS \mid MZ_{MS}, E_T)$] may need to be converted into an adjusted rate [i.e., $P(MS \mid IG_{MS}, E_T)$] because the observed rate may reflect, in part, the fact that *MZ*-twin *probands* share both their intrauterine and <u>some</u> of their other environments with their *co-twin*. If this *co-twin* either has, or will subsequently develop, MS then, potentially, these shared environmental experiences may also make MS more likely in the *proband*. In this case, to isolate the genetic contribution, the impact of these environmental similarities needs to be removed (*see Section 2a*).

By definition, an *exposure-level* can never be greater than its *maximum* value so that:

$[P(E \mid MZ_{MS})_2 \leq 1]$.

Moreover, if *any* susceptible *MZ-proband* $(G_i)$ is known to have experienced $\{E_i\}$, then both the environmental experience of their *co-twin*, and the *Time Period*, become irrelevant such that:

$$P(MS \mid E, MZ_{MS})_2 = P(MS \mid E)_2 = P(MS \mid E)$$

Therefore: $$P(MS \mid MZ_{MS})_2 = P(MS, E \mid MZ_{MS})_2 = P(MS \mid E, MZ_{MS})_2 * P(E \mid MZ_{MS})_2$$

or: $$P(MS \mid MZ_{MS})_2 = P(MS \mid E) * P(E \mid MZ_{MS})_2$$

so that: $$P(MS \mid E) \geq P(MS \mid MZ_{MS})_2$$ *Equation S8i*

Thus, the value of the parameter $[P(MS \mid E)]$ must be, at least, as large as the *currently* observed *MZ*-twin concordance rate. And similarly:

$$\boldsymbol{c} = P(MS \mid E, M) \geq P(MS \mid M, MZ_{MS})_2$$ *Equation S8j*

and: $$\boldsymbol{d} = P(MS \mid E, F) \geq P(MS \mid F, MZ_{MS})_2$$ *Equation S8k*

**Table S1a.** Definitions for Terms used in the Mathematical Development – *see also Table 1; Main Text*

| Terms | Definitions |
|---|---|
| $(Z)$ | The population – a set consisting of $(N)$ individuals – *see Main Text* |
| $(F)$ , $(M)$ | Subsets of *women* $(F)$ and *men* $(M)$ within $(Z)$ – *see Main Text* |
| $(MS)$ | Subset of *all* individuals within $(Z)$ who either have, or will subsequently develop, MS – *see Main Text* |
| $(G)$ | Subset of individuals within $(Z)$ who have <u>any</u> non-zero *life-time* chance of developing MS under <u>some</u> environmental conditions – *see Main Text & Section 1a* |
| $G_i$ | The unique genotype of the *i*<sup>th</sup> susceptible individual within $(G)$: $(i = 1,2,\ldots,m)$ – *see Main Text* |
| $p$ | Proportion of *women* in the $(G)$ subset – i.e., $p = P(F \mid G)$ – *see Main Text* |
| $(E_T)$ | Environmental conditions of *some* specific *Time-Period* – *see legend; Table 2; Main Text* |
| *Subscripts (1) & (2)* | Designations for *parameter-values* during *Time Period #1* (*1941-1945*) and *Time Period #2* (*1976-1980*) – e.g., $P(MS)_2$ represents $P(MS)$ during *Time Period #2* – *see Section 5b* |
| $P(MS \mid E_T)$ | *Penetrance* of MS for the population $(Z)$ during $(E_T)$ – *see Main Text* |
| $x = P(MS \mid G, E_T)$ | *Penetrance* of MS for the $(G)$ subset of the population $(Z)$ during $(E_T)$ – *see Main Text* |
| $x_i = P(MS \mid G_i, E_T)$ | *Penetrance* of MS for the *i*<sup>th</sup> individual in the $(G)$ subset of $(Z)$ during $(E_T)$ – *see Main Text* By the definition of $(G)$ – *above* – it <u>must</u> be that, during <u>some</u> $(E_T)$: $\forall\, G_i \in (G)$: $x_i > 0$ |
| $Zw = z_w$ | *Penetrance* of MS for the subset of *susceptible women* $(F, G)$ within $(Z)$ during $(E_T)$ – Also called the *failure probability* for *susceptible women* during $(E_T)$ – *see Sections 3a & 5b* |
| $Zm = z_m$ | *Penetrance* of MS for the subset of *susceptible men* $(M, G)$ within $(Z)$ during $(E_T)$ – Also called the *failure probability* for *susceptible men* during $(E_T)$ – *see Sections 3a & 5b* |
| $\boldsymbol{c}$ , $\boldsymbol{d}$ | Limiting values (constants) for the <u>maximum</u> *failure probability* in *susceptible-men* $(\boldsymbol{c})$; and *susceptible-women* $(\boldsymbol{d})$ – i.e., $(Zm \le \boldsymbol{c} \le 1)$ and $(Zw \le \boldsymbol{d} \le 1)$ – *see Sections 5b-c* |
| $s_a$ , $s_{aw}$ , $s_{am}$ | The ratio of: $[P(MS \mid DZ_{MS})/P(MS \mid S_{MS})]$ ; used to adjust the *MZ*-twin concordance for the environments shared by *MZ*-twins; considered collectively $(s_a)$, or the comparable ratios for *women* $(s_{aw})$ and *men* $(s_{am})$; considered separately – *see Main Text & Sections 2b-c* |
| $x'$, $z'_w$ , $z'_m$ | Adjusted *MS-penetrance* values for members of the $(G, MZ_{MS})$ subset, $(x')$, and for the subsets $(G, F, MZ_{MS})$ – $(z'_w)$ – and $(G, M, MZ_{MS})$ – $(z'_m)$ – considered separately e.g., $x' = P(MS \mid MZ_{MS})/s_a = P(MS \mid IG_{MS})$ – *see Sections 2a, 3a & 4a-b* |
| $(X)$ | Set of *MS-penetrance* values for *all* $(m)$ members of the "*genetically-susceptible*" subset $(G)$ – i.e., $(X) = (x_1, x_2, \ldots, x_m)$ – *see Main Text & Section 4a* |
| $\sigma_X^2$ , $\sigma_w^2$ , $\sigma_m^2$ | Variance of the *MS-penetrance* values for *all* susceptible individuals $(\sigma_X^2)$ and for susceptible *women*, $(\sigma_w^2)$, and *men*, $(\sigma_m^2)$, considered separately – *see Sections 3a & 4a* |
| $(G_w)$ , $(G_m)$ | Alternative designations for subsets of all susceptible *women* – i.e., $(G_w) = (F, G)$ – and all susceptible *men* – i.e., $(G_m) = (M, G)$ – *see Sections 3a, 4a & 7h* |
| $G_{dw}$ , $G_{dm}$ | Alternative designations for the genotypes of the $(mp)$ *women* in the $(F, G)$ subset – $(d = 1,2,\ldots,mp)$ – and for the genotypes of the $[m(1-p)]$ *men* in the $(M, G)$ subset – $[d = 1,2,\ldots,m(1-p)]$ – *see Sections 3a, 4a & 7h* |
| $z_{dw}$ , $z_{dm}$ | *MS-Penetrance* values for the *d*<sup>th</sup> *susceptible woman* in $(G_w)$: $(d = 1,2,\ldots,mp)$ ; and for the *d*<sup>th</sup> *susceptible man* in $(G_m)$: $[d = 1,2,\ldots,m(1-p)]$ – *see Section 3a* |

**Table S1b.** Definitions for Terms used in the Mathematical Development – *Continued*

| **Terms** | **Definitions** |
|---|---|
| $G_{i1}$ , $G_{i2}$ | Any pair of *susceptible* individuals, randomly-selected from $(G)$ – *see Section 3a* |
| $x_{i1}$ , $x_{i2}$ | *MS-penetrance* values, respectively, for the individuals $G_{i1}$ and $G_{i2}$ – *see Section 3a* |
| $(G_{is})$ | Subset of susceptible individuals who share the same "*susceptibility genotype*" with the *i*$^{th}$ susceptible individual – i.e., the genotype considering *only* those genetic factors related to "*genetic susceptibility*" – *see Sections 7f-h* |
| $(G_s)$ | Subset of all "*susceptibility genotypes*" within $(Z)$ – *see Sections 7f-h* |
| $(G_{ia})$ | Subset of susceptible individuals who share the same "*autosomal susceptibility genotype*" with the *i*$^{th}$ susceptible individual – i.e., the genotype considering *only* those *autosomal* genetic factors related to "*genetic susceptibility*" – *see Sections 7f-h* |
| $(G_a)$ | Subset of all "*autosomal susceptibility genotypes*" within $(Z)$ – *see Sections 7f-h* |
| $(G_{it})$ | Subset of susceptible individuals (possibly with different *susceptibility genotypes*) who are in the same "*i-type*" *exposure-group* – i.e., individuals who share the same $\{E_i\}$ family of "*sufficient*" *environmental-exposures* – *see Sections 7f-h* |
| $\{G_t\}$ | Family of all "*i-type*" *exposure-groups* within $(Z)$ – *see Sections 7f-h* |
| $\{E_i\}$ | Family of <u>*every*</u> set of *environmental-exposures*, each of which is "*sufficient*", by itself, to *cause* MS in the *i*$^{th}$ susceptible individual within $(G)$: $(i = 1,2, \ldots, m)$ – *see Section 1a* |
| $(E)$ | Event that a randomly selected member of $(G)$ – the *proband* – experiences an environment *sufficient* to *cause* MS in them – *see Section 1a* |
| $P(E \mid G, E_T)$ | Probability that the event $(E)$ occurs during $(E_T)$ – *see Section 1a* |
| $u$ | Variable representing the level of *environmental-exposure*, as measured by the odds that the event $(E)$ occurs during *any* $(E_T)$ – *see Section 5a* |
| $a$ | Level of *environmental-exposure* during *some* specific $(E_T)$ – i.e., when: $(u = a)$ |
| $h(u)$ , $k(u)$ | Unknown (and unspecified) hazard functions for *susceptible-men* – $h(u)$ ; and for *susceptible-women* – $k(u)$ – *see Section 5a* |
| $H(a)$ , $K(a)$ | Cumulative hazard functions for *susceptible-men* – $H(a)$ ; and *susceptible-women* – $K(a)$ – Defined as the definite integrals of these unknown and unspecified hazard functions from an *exposure-level* of: $(u = 0)$ to an *exposure-level* of: $(u = a)$ – *see Section 5a* |
| $R > 0$ | Value of the proportionality-factor (*if the hazards are proportional*) – i.e., $k(u) = R * h(u)$ – *see Main Text & Sections 5a & 7a* |
| $R^{app}$ | The "apparent" value of $R$ – i.e., the value of $R$ for proportional hazards when: $(\boldsymbol{c} = \boldsymbol{d} \leq 1)$ – *see Section 7b* |
| $C$ | Ratio of the *MS-penetrance* during *Time Period #1*, $[P(MS)_1]$, to that during *Time Period #2*, $[P(MS)_2]$ – i.e., $C = [P(MS)_1/P(MS)_2]$ – *see Section 8a* |
| $C_F$ , $C_M$ | Analogous ratios to $(C)$ considering *women* $(C_F)$ and *men* $(C_M)$ separtately. – i.e., $C_F = [P(MS,F)_1/P(MS,F)_2]$ & $C_M = [P(MS,M)_1/P(MS,M)_2]$ – *see Section 8a* |
| $\lambda_w$ , $\lambda_m$ | *Environmental-exposure* thresholds for developing MS in susceptible *women* $(\lambda_w)$ and susceptible *men* $(\lambda_m)$ – *see Main Text & Section 5c* |
| $\lambda$ | Difference in the *environmental-exposure* threshold between susceptible *women* and susceptible *men* – i.e., $\lambda = \lambda_w - \lambda_m$ – *see Main Text & Section 5c* |

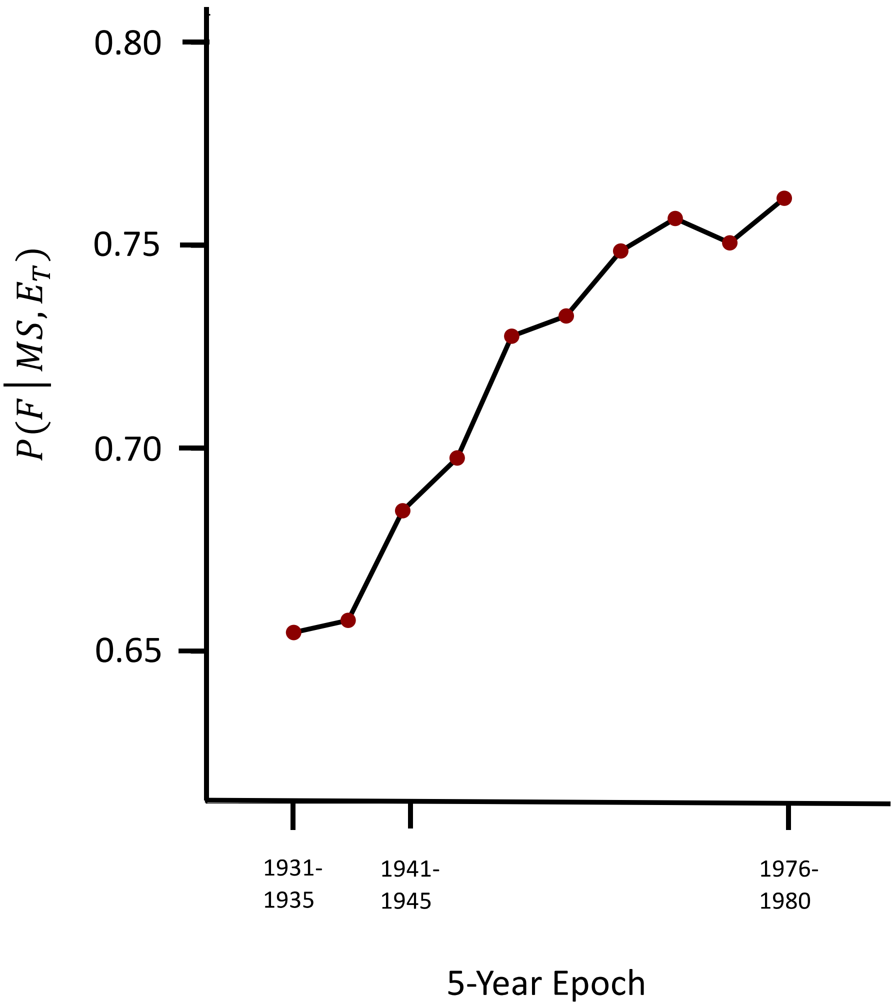

**Figure S1.** Reported change [23] in the proportion of *women* among MS patients (*y-axis*) – i.e., $[P(F \mid MS, E_T)]$ – over the time of birth-year date (*x-axis)* for persons born in Canada from 1931 until 1980. Each data point in the *Figure* represents a sequential 5-year epoch beginning with $(1931 - 1935)$ and ending with $(1976 - 1980)$. In the *CCPGSMS* dataset there were total of $(29{,}748)$ identified *MS-cases*, of whom $(27{,}074)$ were born during this *date-range* and who were included in the analysis [23]. Each 5-year epoch from $(1931 - 1980)$ contained a minimum of $(500)$ identified patients and, of the total number of patients identified in this *date-range*, $(19{,}417)$ were *women* and $(7{,}657)$ were *men*. In addition, there were an average of $(2{,}400)$ patients identified in each 5-year epoch, for an average of $(480)$ patients in each birth-year [23]. For purposes of the present analysis, the epoch of $(1941 - 1945)$ was chosen as *Time Period #1* because it was the earliest epoch with a very small *confidence-interval* [23]. The epoch of $(1976 - 1980)$ was chosen as *Time Period #2* because it represents the most recent of the reported Canadian epochs [23]. Nevertheless, choosing *any* 5-year epoch from $(1931 - 1975)$ as *Time Period #1* still demonstrates and increasing proportion of *women* between *Time Period #1* and *Time Period #2*.

**Table S2.** Epidemiological Data regarding Multiple Sclerosis in Canada circa (*2000 –2015*)

| ***Canadian Data for MZ Twin-Pairs**** | ***Women*** | ***Men*** | ***Totals*** |
|---|---|---|---|
| Concordant for MS | 22 | 2 | 24 |
| Discordant for MS | 66 | 43 | 109 |
| Totals | 88 | 45 | 133 |
| *Proband-wise Concordance*** | 0.340 | 0.065 | 0.253 |
| *Proportion Concordant* | 0.917 | 0.083 | 1.000 |
| *Proportion Discordant* | 0.606 | 0.394 | 1.000 |

***Population Data for Canada in 2001-2010:***

$Total\ Population = 34{,}108{,}800\ \ individuals$ -- *from the 2010 Canadian census* [24]

$P(F) = 0.504$ -- *from the 2010 Canadian census* [24]

*MS-prevalence* (*~2001*) $= (100 - 128)\ cases\ per\ (100{,}000)\ population$ -- *from Reference* [5]

***Case Ascertainment in the CCPGSMS:***

*Estimated using a Twin-rate* $= (0.0091)$ *Twins per birth, and an MS-prevalence* $= 0.001$*, in Canada*

-- $(454)\ indetified\ cases\ /\ (547)\ expected\ cases = 83.0\%$ -- *from Reference* [5]

***Summary Data for MS-Concordance among DZ Twins and Non-twin Siblings in Canada:***

$P(MS \mid DZ_{MS}) = 0.054$ -- *from Reference* [5]

$P(MS \mid S_{MS}) = 0.029$ -- *from Reference* [5]

***Summary Data for the Preponderance of Women among MS Patients in Canada***

$P(F \mid MS) = 19{,}417/27{,}074 = 0.717$ -- *from Reference* [23]

$P(F \mid MS, MZ_{MS}) = 22/24 = 0.917$ -- *from Table, above, Reference* [5]

*During Time Period #1* (*1941-1945*): $P(F \mid MS) = 0.685$ -- *from Figure S1 (above); Reference* [23]

*During Time Period #2* (*1976-1980*): $P(F \mid MS) = 0.762$ -- *from Figure S1 (above); Reference* [23]

* Data drawn from the MS-patients in the *CCPGSMS* database as of (~2001) – *Reference* [5]

** *Proband-wise* (or *case-wise*) concordance calculated according to [5,25] -- adjusted for double ascertainments $(13/24 = 54\%)$

-- *Proband-wise Concordance* in *men* $= P(MS \mid M, MZ_{MS})$

-- *Proband-wise Concordance* in *women* $= P(MS \mid F, MZ_{MS})$
